# Ethnic and Social Health Inequalities in Body Mass Index Trajectories through Childhood and Adolescence: A Longitudinal Population-Based Study in Leicestershire UK

**DOI:** 10.64898/2026.04.15.26350938

**Authors:** Lorenz M. Leuenberger, Fabiën N. Belle, Ben D. Spycher, Myrofora Goutaki, David K.H. Lo, Erol A. Gaillard, Claudia E. Kuehni

**Affiliations:** Institute of Social and Preventive Medicine, University of Bern, Switzerland; Graduate School for Health Sciences, University of Bern, Switzerland; Department of Paediatrics, Inselspital, Bern University Hospital, University of Bern, Bern, Switzerland; Department of Respiratory Sciences, Leicester NIHR Biomedical Research Centre (Respiratory theme), University of Leicester, Leicester, UK; Department of Paediatric Respiratory Medicine, Leicester Children’s Hospital, University Hospitals Leicester, Leicester, UK

## Abstract

**Background:** Ethnic minorities and socioeconomically disadvantaged populations in the UK are at increased risk of obesity. We modelled longitudinal body mass index (BMI) trajectories through infancy, childhood, and adolescence to identify at-risk groups and modifiable risk factors.

**Methods:** This cohort sampled 10,350 White and South Asian children born in Leicestershire, 1985–1997. We included 5,571 participants with ≥3 BMI measurements between 0–18 years collected from healthcare records, questionnaires, and study visits. We used Group-Based Trajectory Modelling of BMI, separately by sex and ethnicity, and combined. We identified at-risk groups and modifiable risk factors using multinomial logistic regression, with inverse probability weighting to reduce selection bias.

**Results:** We identified similar five BMI trajectories across sex and ethnicity: stable normal BMI (47%); persistent low BMI (30%); early overweight resolving (8%); childhood onset obesity (4%); and adolescent onset overweight (11%). Childhood onset obesity deviated from stable normal BMI at 2–4 years of age, adolescent onset overweight at 4–6 years. South Asians were at higher risk of childhood onset obesity (aOR: 1.66 [95%CI 1.08–2.53]) and adolescent onset overweight (1.29 [0.98–1.71]) than Whites. Children from deprived backgrounds (1.66 [0.92–2.82], most vs least deprived quintile) and those with less educated parents (1.67 [1.08–2.63], compulsory vs higher education) were at increased risk of childhood onset obesity. Smoking during pregnancy (1.50 [0.88–2.54]) and absence of breastfeeding (1.56 [1.07–2.29]) increased risk of childhood onset obesity. Physical activity decreased risk of childhood onset obesity (0.64 [0.44–0.93], ≥4 vs 0–3 hours/week) and adolescent onset overweight (0.75 [0.59–0.94]).

**Conclusion:** BMI trajectories diverge as early as age 2 years, revealing ethnic and social inequalities. Obesity strategies in the UK should intervene during critical windows in early life and prioritise South Asian children and those from socioeconomically deprived backgrounds.

## INTRODUCTION

In the United Kingdom, obesity is unequally distributed across socioeconomic and ethnic lines. For example, national reports show higher obesity prevalence in children from ethnic minorities and socioeconomically deprived areas. ^1,2^ Socioeconomic inequalities in body mass index (BMI) have increased over recent decades and appeared to widen during adolescence. ^3,4^ The consequences of childhood obesity are profound, including increased risks of lifelong obesity, and later cardiovascular disease, cancer, and infectious diseases. ^5,6^

Most evidence regarding these disparities is based on cross-sectional data, which identifies groups disproportionately affected by obesity, but cannot reveal how obesity develops over time. For this, we need longitudinal studies of BMI trajectories, which help to understand when differences in BMI begin and allow to identify critical windows for interventions and for better targeting prevention strategies. Such studies are rare and research that focuses on ethnic minorities and socioeconomically deprived populations is even less common. In 2019, a systematic review identified 14 studies that had modelled BMI trajectories from birth. ^7^ Half of them were limited in sample size (N<1,500) and only one assessed BMI beyond childhood (>12 years). Two recent studies described BMI trajectories from birth to early and mid-adulthood indicating that obesity may develop early in life, but they did not investigate ethnic minorities separately. ^8,9^ A UK study found lower mean BMI from 3–14 years in South Asian compared to White children, but did not investigate how overweight or obesity trajectories develop. ^10^ Consequently, evidence on obesity in South Asian children remains inconclusive, ^11^ as population-level averages may mask high-risk subgroups.

To address this gap, we analysed distinct BMI trajectories from early childhood through adolescence in a socioeconomically diverse UK cohort including White and South Asian children. We modelled BMI trajectories separately for White and South Asian children, and combined, to determine critical windows. We investigated associations of demographic, socioeconomic, perinatal, and lifestyle factors with overweight and obesity trajectories to identify at-risk groups and risk factors.

## MATERIALS AND METHODS

### Study design and data collection

We analysed data from the Leicester Respiratory Cohorts, population-based childhood cohort studies that investigate respiratory health; ^12–15^ their design and procedures were previously described. ^16^ Two random samples were drawn of children born in Leicestershire, UK: the 1990 cohort (1,650 White children, born 1985–1989) and 1998 cohort (8,700 White and South Asian children, born 1993–1997). Birth records and information on well-child visits came from the Leicester Health Authority Child Health Database. Families completed up to six postal questionnaires (1990–2010) during infancy, childhood, and adolescence to assess growth, development, and respiratory outcomes, and cover socioeconomic, perinatal, and lifestyle factors. Most children also had a clinical study visit. ^16^ The Leicestershire Health Authority Research Ethics Committee approved the study (Ref numbers 4867, 5005, 04/Q2501/61, 07/H0407/70), and written consent was obtained from children and parents. See eSupplement-Table 1 for more information on data collection.

**Table 1:**
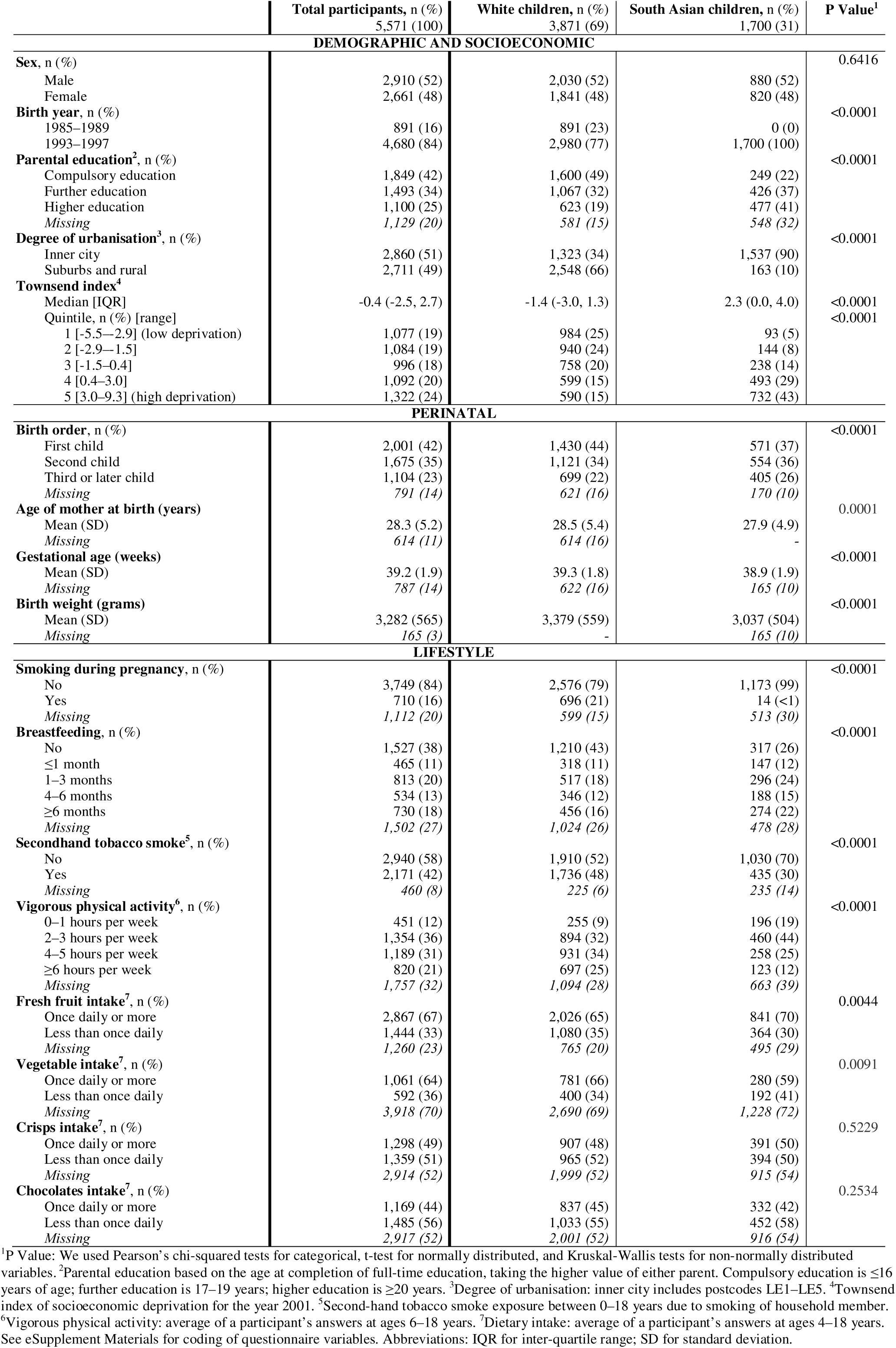
Demographic, socioeconomic, perinatal, and lifestyle characteristics of the participants in the Leicester Respiratory Cohorts included in the BMI trajectory analysis.

### Outcome: Body mass index

Information on height and weight included measurements from well-child visits, the clinical study visit, and parent-reported or self-reported values from questionnaires. We identified and corrected unit errors (e.g., inches instead of centimetres) and switched recordings (e.g. height recorded as weight) based on height and weight z-scores calculated with UK-WHO growth charts. We manually checked and corrected outliers the R package *growthcleanr* identified. We defined BMI as weight/height^2^, calculated UK-WHO z-scores, and excluded BMI values at ≥18 years and implausible BMI values (z-score <-5 or >5). ^8^ We included participants with at least 3 BMI values from 0–18 years, of which one must have been between 0–1 years and one between 3–10 years.

### Ethnicity

Ethnicity was self-reported in birth records; mothers self-identified as Bangladeshi, Pakistani, Indian, or any other Asian background. Leicestershire saw initial migration of predominantly Indian and Pakistani communities after the Partition of the British Empire in India in 1947. ^17^ Subsequent migration of East African Asians came in the 1970s after several African countries gained independence including Kenia and Uganda. ^17^ By the 1980s, the South Asian demographic in Leicestershire diversified with migration from Bangladeshi communities. ^17^

### Socioeconomic, perinatal and lifestyle factors

We considered socioeconomic, perinatal, and lifestyle factors as exposures based on previous literature and availability in the study. ^3,4,7–10^ Socioeconomic variables included individual-level parental education from questionnaires, area-based degree of urbanization, and Townsend index of socioeconomic deprivation linked to the post code of the family home at recruitment. We defined the post codes LE1 to LE5 as “inner city” and other postcodes as “suburbs or rural”. We used the continuous Townsend index and its quintiles from the 2001 UK-wide census. Perinatal variables included maternal age at birth, birth order, gestational age, and birth weight from birth records. Lifestyle variables included questionnaire-reported cigarette smoking during pregnancy, duration of breastfeeding, exposure to secondhand tobacco smoke of other household members, number of hours per week spent on vigorous physical activity, and detailed dietary intake (fresh fruit, vegetables, crisps, sweets). We defined exposure to secondhand tobacco smoke if reported on one or more occasions. For participants with multiple questionnaire answers, we averaged physical activity and dietary intake across questionnaires from ages 6–18 and 4–18 years. The questionnaire and coded variables used for this analysis are shown in eSupplement-Table 2.

**Table 2:** Demographic, socioeconomic, perinatal, and lifestyle characteristics of study participants of the Leicester Respiratory Cohorts stratified by BMI trajectories.

|  | Stable normal<br>BMI, n (%) | Persistent low<br>BMI, n (%) | Early overweight<br>resolving, n (%) | Childhood onset<br>obesity, n (%) | Adolescent onset<br>overweight, n (%) | P Value<sup>1</sup> |
| --- | --- | --- | --- | --- | --- | --- |
|  | 2,625 (47) | 1,648 (30) | 442 (8) | 235 (4) | 621 (11) |  |
| DEMOGRAPHIC AND SOCIOECONOMIC |  |  |  |  |  |  |
| Sex, n (%) |  |  |  |  |  |  |
| Male | 1,505 (57) | 742 (45) | 252 (57) | 120 (51) | 291 (47) | <0.0001 |
| Female | 1,120 (43) | 906 (55) | 190 (43) | 115 (49) | 330 (53) |  |
| Birth year, n (%) |  |  |  |  |  |  |
| 1985–1989 | 459 (17) | 211 (13) | 88 (20) | 35 (15) | 98 (16) | 0.0002 |
| 1993–1997 | 2,166 (83) | 1,437 (87) | 354 (80) | 200 (85) | 523 (84) |  |
| Ethnicity, n (%) |  |  |  |  |  |  |
| White | 2,026 (77) | 913 (55) | 361 (82) | 156 (66) | 415 (67) | <0.0001 |
| South Asian | 599 (23) | 735 (45) | 81 (18) | 79 (34) | 206 (33) |  |
| Parental education<sup>2</sup>, n (%) |  |  |  |  |  |  |
| Compulsory education | 916 (44) | 471 (38) | 161 (44) | 89 (46) | 212 (40) | 0.0100 |
| Further education | 707 (34) | 436 (35) | 115 (32) | 65 (34) | 170 (32) |  |
| Higher education | 482 (23) | 346 (28) | 86 (24) | 40 (21) | 146 (28) |  |
| Missing | 520 (20) | 395 (24) | 80 (18) | 41 (17) | 93 (15) |  |
| Degree of urbanisation<sup>3</sup>, n (%) |  |  |  |  |  |  |
| Inner city | 1,196 (46) | 984 (60) | 193 (44) | 137 (58) | 350 (56) | <0.0001 |
| Suburbs and rural | 1,429 (54) | 664 (40) | 249 (56) | 98 (42) | 271 (44) |  |
| Townsend index<sup>4</sup> |  |  |  |  |  |  |
| Median [IQR] | -0.7 (-2.7, 2.4) | 0.5 (-2.2, 3.3) | -1.0 (-2.8, 1.4) | 0.5 (-2.1, 3.2) | -0.3 (-2.5, 2.9) | <0.0001 |
| Quintile, n (%) |  |  |  |  |  |  |
| 1 (low deprivation) | 566 (22) | 268 (16) | 97 (22) | 31 (13) | 115 (19) | <0.0001 |
| 2 | 546 (21) | 277 (17) | 91 (21) | 46 (20) | 124 (20) |  |
| 3 | 470 (18) | 275 (17) | 108 (24) | 41 (17) | 102 (16) |  |
| 4 | 476 (18) | 373 (23) | 62 (14) | 54 (23) | 127 (20) |  |
| 5 (high deprivation) | 567 (22) | 455 (28) | 84 (19) | 63 (27) | 153 (25) |  |
| PERINATAL |  |  |  |  |  |  |
| Birth order, n (%) |  |  |  |  |  |  |
| First child | 932 (41) | 603 (43) | 152 (41) | 73 (36) | 241 (45) | 0.2579 |
| Second child | 810 (36) | 485 (34) | 119 (32) | 84 (41) | 177 (33) |  |
| Third or later child | 517 (23) | 325 (23) | 98 (27) | 47 (23) | 117 (22) |  |
| Missing | 366 (14) | 235 (14) | 73 (17) | 31 (13) | 86 (14) |  |
| Age of mother at conception (years) |  |  |  |  |  |  |
| Mean (SD) | 28.3 (5.3) | 28.1 (5.2) | 28.7 (5.6) | 29.1 (5.0) | 28.7 (5.0) | 0.0090 |
| Missing | 310 (12) | 148 (9) | 62 (14) | 25 (11) | 69 (11) |  |
| Gestational age (weeks) |  |  |  |  |  |  |
| Mean (SD) | 39.3 (1.8) | 39.0 (2.0) | 39.5 (1.5) | 39.2 (1.9) | 39.1 (1.8) | <0.0001 |
| Missing | 366 (14) | 230 (14) | 73 (17) | 32 (14) | 86 (14) |  |
| Birth weight (grams) |  |  |  |  |  |  |
| Mean (SD) | 3,359 (545) | 3,094 (565) | 3,563 (498) | 3,374 (561) | 3,205 (540) | <0.0001 |
| Missing | 54 (2) | 80 (5) | 10 (2) | 6 (3) | 15 (2) |  |
| LIFESTYLE |  |  |  |  |  |  |
| Smoking during pregnancy, n (%) |  |  |  |  |  |  |
| No | 1,743 (83) | 1,130 (90) | 285 (79) | 148 (76) | 443 (84) | <0.0001 |
| Yes | 369 (17) | 131 (10) | 76 (21) | 48 (24) | 86 (16) |  |
| Missing | 513 (20) | 387 (23) | 81 (18) | 39 (17) | 92 (15) |  |
| Breastfeeding, n (%) |  |  |  |  |  |  |
| No | 723 (38) | 422 (35) | 123 (38) | 81 (45) | 178 (37) | 0.1301 |
| ≤1 month | 229 (12) | 125 (10) | 44 (14) | 17 (9) | 50 (10) |  |
| 1–3 months | 382 (20) | 247 (21) | 59 (18) | 37 (21) | 88 (18) |  |
| 4–6 months | 240 (13) | 171 (14) | 41 (13) | 14 (8) | 68 (14) |  |
| ≥6 months | 311 (16) | 229 (19) | 58 (18) | 30 (17) | 102 (21) |  |
| Missing | 740 (28) | 454 (28) | 117 (26) | 56 (24) | 135 (22) |  |
| Secondhand tobacco smoke<sup>5</sup>, n (%) |  |  |  |  |  |  |
| No | 1,349 (56) | 934 (63) | 208 (50) | 116 (51) | 333 (56) | <0.0001 |
| Yes | 1,048 (44) | 544 (37) | 205 (50) | 113 (49) | 261 (44) |  |
| Missing | 228 (9) | 170 (10) | 29 (7) | 6 (3) | 27 (4) |  |
| Vigorous physical activity<sup>6</sup>, n (%) |  |  |  |  |  |  |
| 0–1 hours per week | 180 (11) | 146 (14) | 35 (12) | 20 (10) | 70 (13) | 0.0001 |
| 2–3 hours per week | 555 (33) | 383 (36) | 99 (33) | 90 (45) | 227 (41) |  |
| 4–5 hours per week | 566 (33) | 312 (30) | 98 (32) | 53 (27) | 160 (29) |  |
| ≥6 hours per week | 405 (24) | 210 (20) | 72 (24) | 35 (18) | 98 (18) |  |
| Missing | 919 (35) | 597 (36) | 138 (31) | 37 (16) | 66 (11) |  |
| Fresh fruit intake<sup>7</sup>, n (%) |  |  |  |  |  |  |
| Once daily or more | 1,316 (67) | 813 (66) | 217 (63) | 154 (75) | 367 (65) | 0.0310 |
| Less than once daily | 654 (33) | 411 (34) | 130 (37) | 50 (25) | 199 (35) |  |
| Missing | 655 (25) | 424 (26) | 95 (21) | 31 (13) | 55 (9) |  |
| Vegetable intake<sup>7</sup>, n (%) |  |  |  |  |  |  |
| Once daily or more | 481 (65) | 282 (63) | 88 (67) | 41 (62) | 169 (62) | 0.7393 |
| Less than once daily | 254 (35) | 168 (37) | 43 (33) | 25 (38) | 102 (38) |  |
| Missing | 1,890 (72) | 1,198 (73) | 311 (70) | 169 (72) | 350 (56) |  |
| Crisps intake<sup>7</sup>, n (%) |  |  |  |  |  |  |
| Once daily or more | 564 (48) | 358 (50) | 92 (46) | 66 (49) | 218 (51) | 0.5983 |
| Less than once daily | 616 (52) | 357 (50) | 110 (54) | 68 (51) | 208 (49) |  |
| Missing | 1,445 (55) | 933 (57) | 240 (54) | 101 (43) | 195 (31) |  |
| Chocolates intake<sup>7</sup>, n (%) |  |  |  |  |  |  |
| Once daily or more | 522 (44) | 325 (46) | 78 (39) | 60 (45) | 184 (43) | 0.5000 |
| Less than once daily | 659 (56) | 387 (54) | 124 (61) | 73 (55) | 242 (57) |  |
| Missing | 1,444 (55) | 936 (57) | 240 (54) | 102 (43) | 195 (31) |  |

<sup>1</sup>P Value: We used Pearson's chi-squared tests for categorical, one-way ANOVA for normally distributed, and Kruskal-Wallis tests for non-normally distributed variables.
<sup>2</sup>Parental education based on the age at completion of full-time education, taking the higher value of either parent. Compulsory education is ≤16 years of age; further education is 17–19 years of age; higher education is ≥20 years of age. <sup>3</sup>Degree of urbanisation: inner city includes postcodes LE1–LE5. <sup>4</sup>Townsend index of socioeconomic deprivation for the year 2001. <sup>5</sup>Secondhand tobacco smoke exposure between 0–18 years due to smoking of household member. <sup>6</sup>Vigorous physical activity: average of a participant's answers at ages 6–18 years. <sup>7</sup>Dietary intake: average of a participant's answers at ages 4–18 years. See eSupplement-Table 2 for coding of questionnaire variables. Abbreviations: BMI for body mass index; IQR for inter-quartile range; SD for standard deviation

### Statistical analyses

#### Latent BMI trajectory modelling

We modelled distinct BMI development trajectories from 0–18 years using the four step approach of Ram and Grimm, ^18^ see eSupplement methods for detailed description. After literature review, we chose to use Group-Based Trajectory Modelling (GBTM) to identify distinct classes of BMI development. Unlike Growth Mixture Modelling, which includes random effects to model individual variation, GBTM can identify discrete latent classes and group participants. We modelled raw BMI which is a better measure of adiposity change than BMI z-scores. ^19^ We evaluated parametric functions, including polynomials and splines up to the 3^rd^ degree, to flexibly model age patterns of BMI: we performed a grid search on a 1-class model and selected the best function based on model fit (AIC, BIC) and biological plausibility, by visually comparing with the UK-WHO growth charts. Using the selected function, we modelled up to seven BMI classes, first separately by sex and ethnicity, and then combined after visual comparison. We estimated the models with the *lcmm* package in R with 50 random starts and 20 iterations to avoid local maxima solutions. We determined the optimal number of classes based on model fit, class size, entropy level, average posterior probability, proportion of participants with posterior probability ≥0.7, and biological plausibility.

#### Identification of at-risk groups and risk factors for BMI trajectories

We assessed associations of demographic, socioeconomic, perinatal, and lifestyle factors with BMI trajectories using descriptive statistics and univariable multinomial logistic regression. To identify population subgroups with elevated risk (at-risk groups), we used multivariable regression that included demographic, socioeconomic, and perinatal factors. To identify potentially modifiable risk factors, we additionally included lifestyle factors. We also identified risk factors separately by ethnic subgroup.

We accounted for the uncertainty of class membership from the GBTM by weighting each participant’s contribution to the regression model by their posterior probability of membership for each identified trajectory.

Some participants had no recorded measurements, and some questionnaires were only sent to subgroups, resulting in missing data of exposures and the outcome. We evaluated several methods for handling the missing data using DAGs with missingness indicators (eSupplement-Figure 1) and concluded that multiple imputation would likely be biased as exposures and outcome values probably influenced their own missingness (i.e., data were not missing at random).

**Figure 1:**
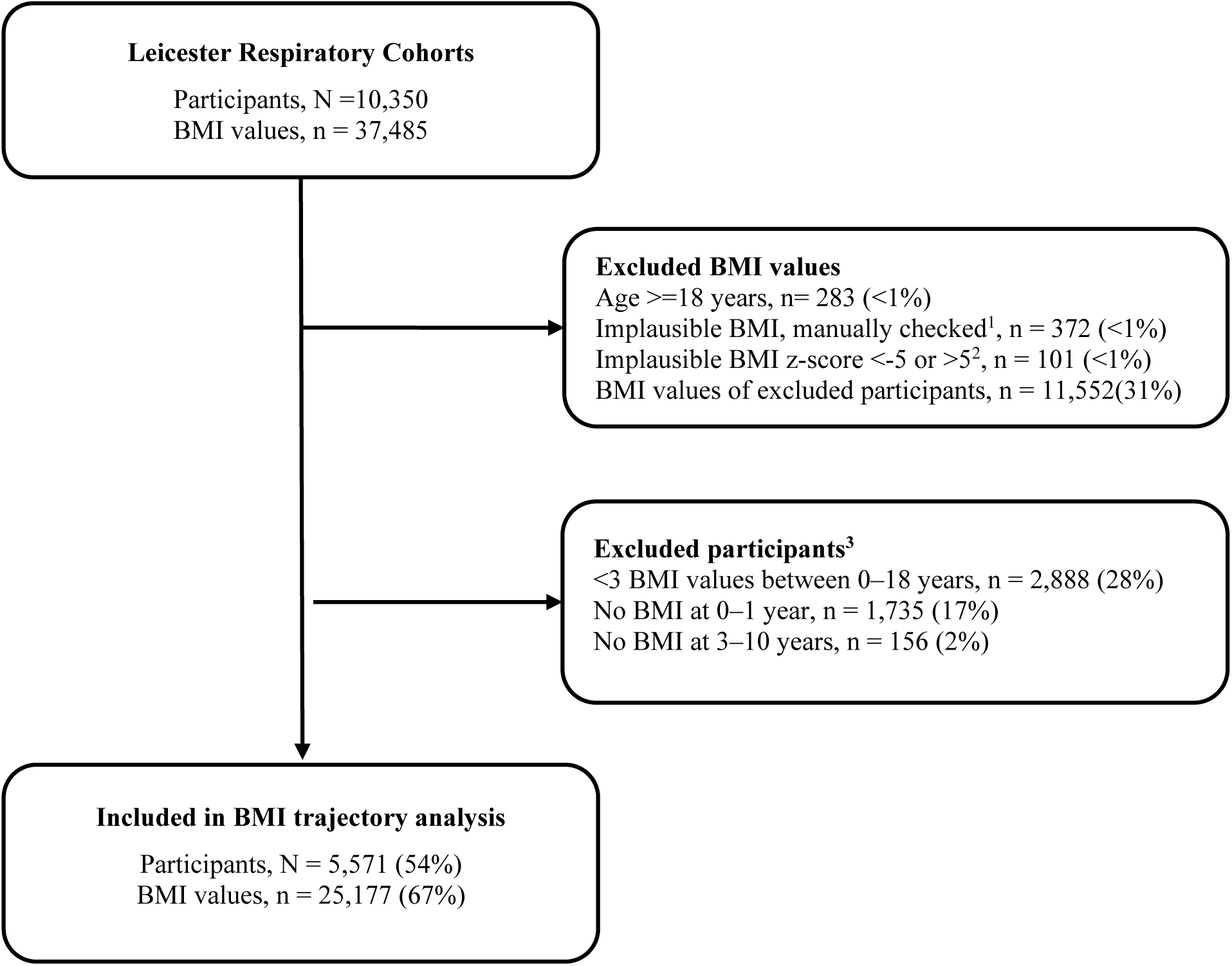
Flow chart of participants of the Leicester Respiratory Cohorts included in the BMI trajectory analysis. ^1^ We manually checked all BMI measurements flagged by the R package *growthcleanr*. We compared them to the UK-WHO growth charts and excluded implausible height, weight, or BMI. We calculated BMI z-scores based on the UK-WHO growth charts and recentred them to our study population. ^2^ After recentring, we excluded BMI z-scores <-5 or >5. ^3^ Participants had to have at least 3 BMI values for inclusion, of which one was at 0–1 years and one was at 3–10 years. Abbreviations: BMI for body mass index

We therefore used complete records analysis with inverse probability weighting (IPW) to reduce selection bias, which returned asymptotically unbiased estimates of odds ratios. ^20^ We compared included and excluded participants and fitted a multivariable logistic regression model to derive stabilized weights based on demographic and socioeconomic characteristics (eSupplement-Table 3 and 4). We evaluated the IPW by calculating standardized differences, comparing included participants to all participants of the Leicester Respiratory Cohorts before and after IPW (eSupplement-Figure 2). We used the stabilized weights and the posterior probabilities of class membership in weighted multinomial logistic regression analyses. The statistical software R Studio version 2023.3.1 and R version 4.2.0 were used for all analyses. The analysis code is available on GitHub (https://github.com/LorenzLeuenberger/BMI-trajectories_Leicester). We followed STROBE and GRoLTS checklists for reporting.

**Figure 2:**
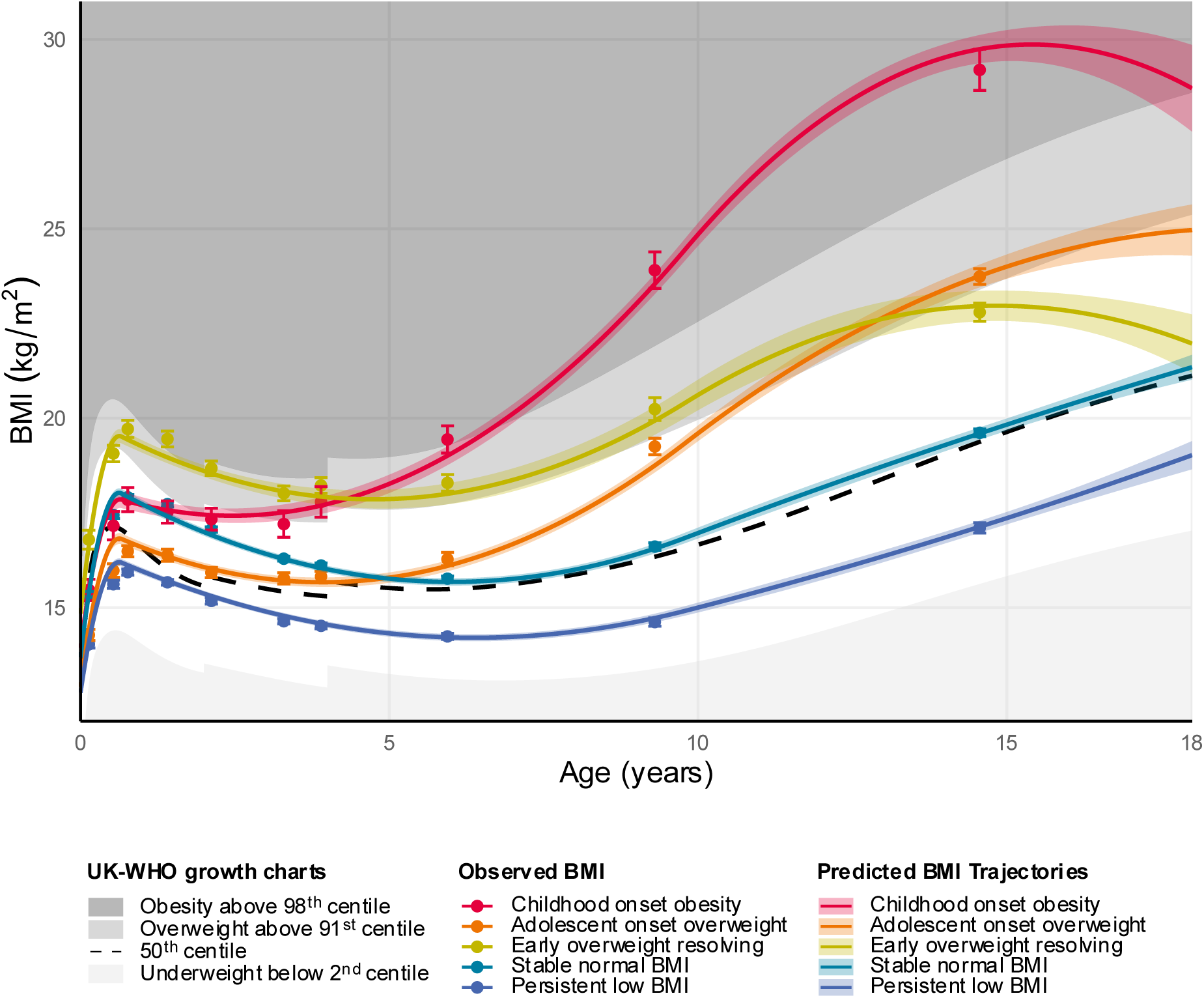
BMI trajectories using Group-Based Trajectory Modelling compared to observed BMI in participants of the Leicester Respiratory Cohorts. We identified five BMI trajectories: *stable normal BMI* (n = 2,625, 47%); *persistent low BMI* (n = 1,648, 30%); *early overweight resolving* (n = 442, 8%); *childhood onset obesity* (n = 235, 4%); *adolescent onset overweight* (n = 621, 11%). The predicted trajectories show model-based marginal means with 95% confidence intervals estimated from the group-based trajectory model. To calculate observed BMI, we divided the dataset into ten age groups with equal numbers of BMI measurements. We assigned participants to the class with their highest posterior probability and calculated the mean and 95% confidence intervals of observed (measured or self-reported) BMI for every age group and class. The plotted centiles are averages of the girl and boy centiles of the UK-WHO growth charts. Abbreviations: BMI for body mass index

## RESULTS

### Study population

The Leicester Respiratory Cohorts sampled 10,350 White and South Asian children. We excluded 2,888 participants who had fewer than three BMI values, 1,735 without BMI between 0–1 years, and 156 without BMI between 3–10 years (Figure 1). We included 5,571 (54%) participants with 25,177 BMI values in the BMI trajectory analysis. Of those, 48% were girls and 69% were White (Table 1). South Asian children had parents with a higher level of education, but lived in more deprived and inner city areas than White children, all p<0.0001. South Asian children were less exposed to maternal smoking during pregnancy and secondhand tobacco smoke of other household members, and were less physically active, all p<0.0001.

Children included in this analysis had a median of 4 [inter quartile range: 4, 5] BMI recordings, most recordings in infancy and childhood (eSupplement-Tables 5 and 6). Based on the 1-class model, we selected a 2^nd^ degree spline function of age with two knots at 0.7 and 9.8 years to model BMI trajectories. We combined all participants in one model because the trajectories of models including only boys, girls, Whites, or South Asians were visually similar (eSupplement-Figure 3). We considered the 5-class model optimal because it had good fit, large enough class sizes, and better entropy and posterior probabilities than the 6- and 7-class models (eSupplement-Table 7 and eSupplement-Figure 4).

**Figure 3:**
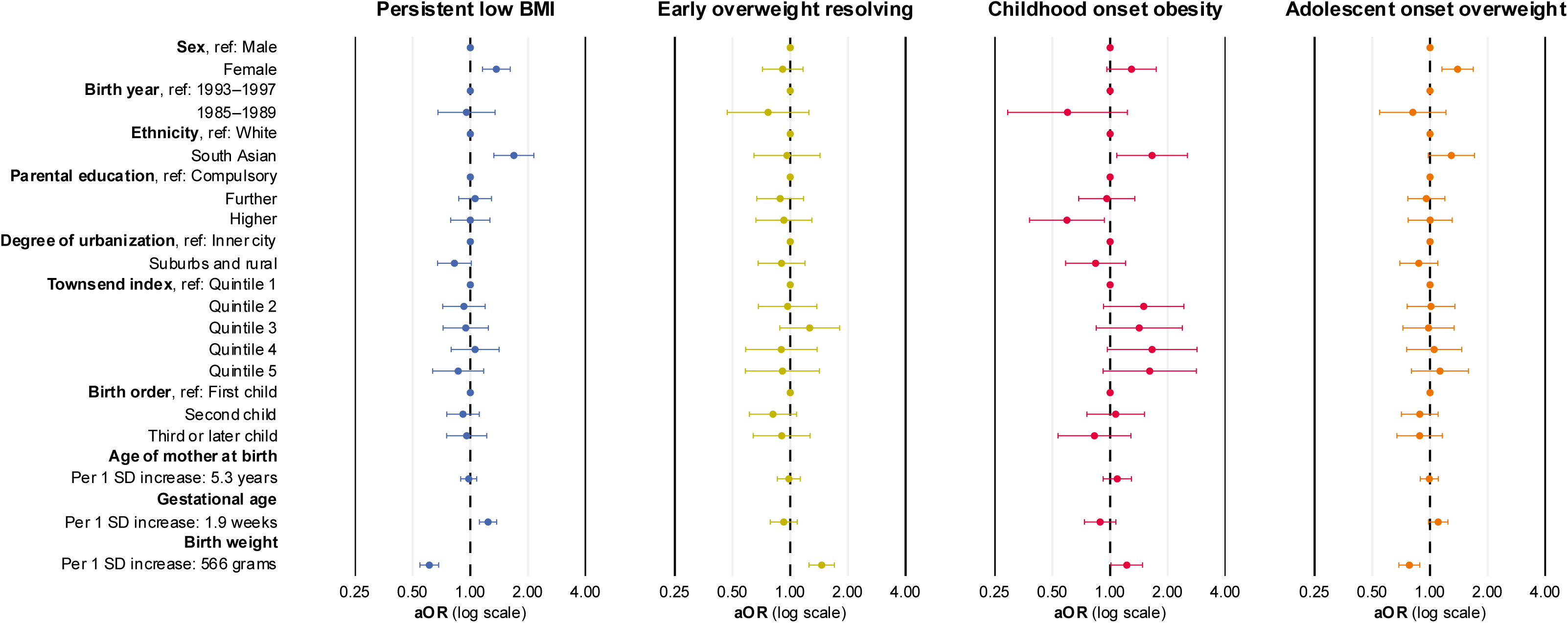
Identification of at-risk groups: Association of demographic, socioeconomic, and perinatal factors with BMI trajectories in participants of the Leicester Respiratory Cohorts. This plot shows adjusted Odds Ratios (aOR) with 95% confidence intervals (CI) from multivariable multinomial logistic regression; the *stable normal BMI* trajectory was the reference. We adjusted the effect of each exposure for all other demographic, socioeconomic, and perinatal factors. This model included n = 3,786 participants, because of data on parental education, birth order, age of mother at birth, gestational age, and birth weight was missing. We took the uncertainty of class membership into account by weighting each participant’s contribution to the regression model by their posterior probability of membership for each of the five BMI trajectories. We used inverse probability weighting to reduce selection bias arising from differences between participants included in the BMI trajectory analysis (vs excluded), see eSupplement Methods. Abbreviations: BMI for body mass index; ref for reference.

**Figure 4:**
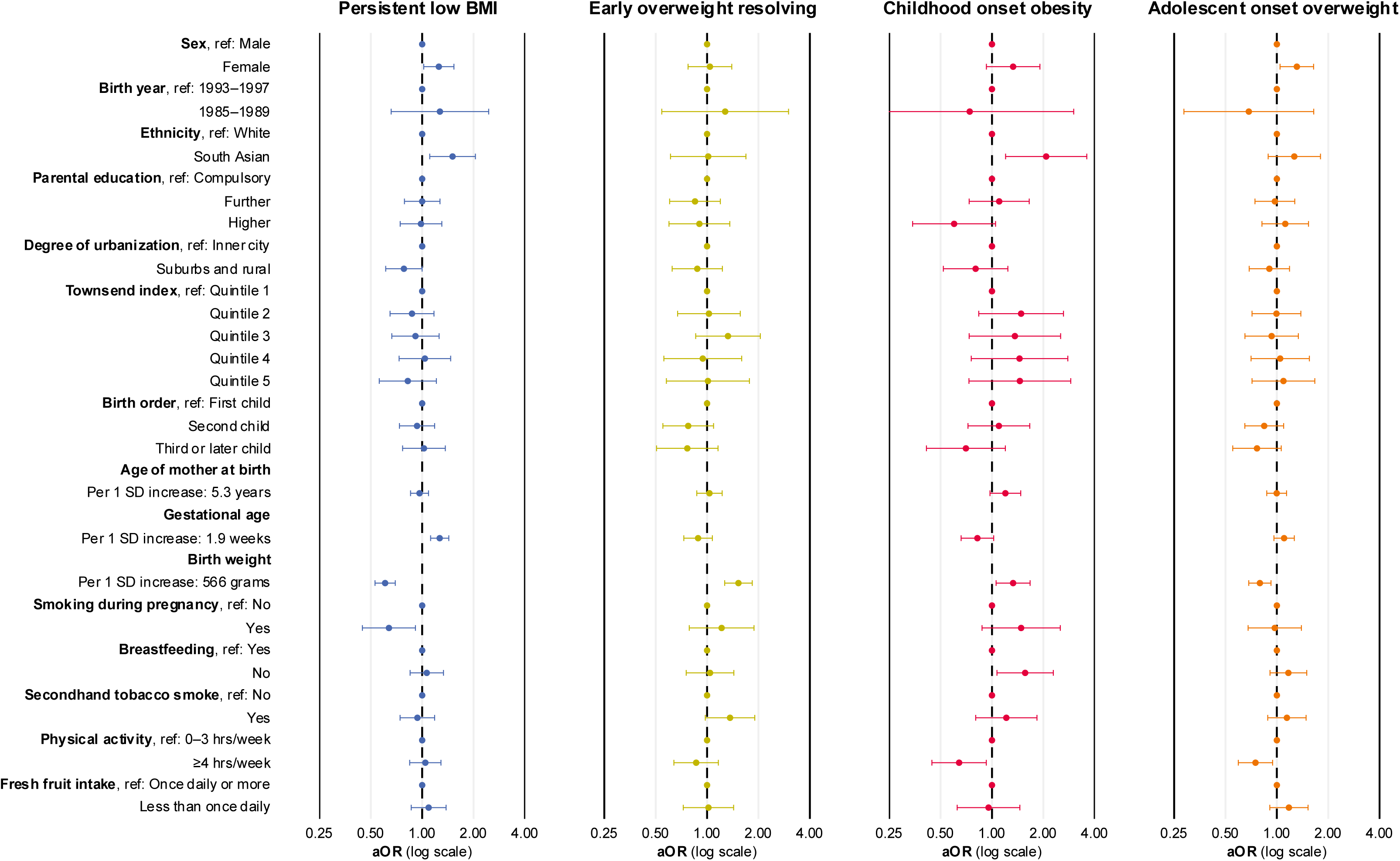
Identification of risk factors: Association of demographic, socioeconomic, perinatal, and lifestyle factors with BMI trajectory in participants of the Leicester Respiratory Cohorts. Adjusted odds ratios are presented with 95% confidence intervals from multinomial logistic regression. We adjusted the effect of each exposure for all other demographic, socioeconomic, perinatal, and lifestyle factors in this table. We did not include all detailed dietary intake variables because of too much data was missing. This model included n = 2,620 participants because data in exposure variables was missing. We recoded breastfeeding and physical activity to avoid small categories. We did not include other detailed dietary intake because >50% of data was missing. We took the uncertainty of class membership into account by weighting each participant’s contribution to the regression model by their posterior probability of membership for each of the five BMI trajectories. We used inverse probability weighting to reduce selection bias arising from differences between participants included in the BMI trajectory analysis (vs excluded), see eSupplement Methods.

### BMI development trajectories

We found five BMI development trajectories (Figure 2): *stable normal BMI* (n = 2,625, 47%); *persistent low BMI* (n = 1,648, 30%); *early overweight resolving* (n = 442, 8%); *childhood onset obesity* (n = 235, 4%); and *adolescent onset overweight* (n = 621, 11%). The mean of the *stable normal BMI* trajectory followed the 50^th^ centile UK-WHO growth charts closely. The *persistent low BMI* trajectory was consistently below the 50^th^ centile, but never dropped below the 2^nd^ centile (underweight). The *early overweight resolving* trajectory was above the 91^st^ centile (overweight) in infancy and childhood; in adolescence, the trajectory resolved towards the 50^th^ centile. *Childhood onset obesity* diverged from *stable normal BMI* between age 2–4 years, became overweight at 5 years, and obese at 7 years. *Adolescent onset overweight* started increasing at age 4–6 years and became overweight at 11 years. Mean predicted BMI trajectories from GBTM aligned well with mean observed BMI values (Figure 2).

### Identification of at-risk groups

Demographic, socioeconomic, and perinatal characteristics differed across the five BMI trajectories (Table 2) and showed associations in univariable regression (eSupplement-Table 8). Compared with *stable normal BMI*, we identified the following at-risk groups for the other four trajectories:

#### Persistent low BMI

When we compared South Asian children (adjusted odds ratio: 1.69 [95% confidence interval: 1.33–2.15]) with White children, and girls (1.37 [1.16–1.62]) with boys the former were more likely to follow the *persistent low BMI* trajectory than the *stable normal BMI* trajectory (Figure 3 and eSupplement-Table 9). Children born at older gestational ages (1.24 [1.12–1.37], per 1.9 weeks) and lower birth weights (1.64 [1.47–1.82], per 565 grams) had higher odds of *persistent low BMI*.

#### Early overweight resolving

Heavier newborns were more likely to follow the *early overweight resolving* trajectory (1.46 [1.25–1.70]).

#### Childhood onset obesity

South Asian children (1.66 [1.08–2.53]), girls (1.29 [0.96–1.74]), those with less educated parents (1.67 [1.08–2.63]; compulsory vs higher education), and those living in socioeconomically deprived areas (1.61 [0.92–2.82]; most vs least deprived quintile) more often had *childhood onset obesity*. Heavier newborns had higher odds of this trajectory (1.22 [1.01–1.48], per 565 grams).

#### Adolescent onset overweight

South Asian children (1.29 [0.98–1.71]) and girls (1.39 [1.15–1.68]) more often had *adolescent onset overweight*. Children born at older gestational age (1.10 [0.98–1.24]) were more likely to follow this trajectory.

### Identification of modifiable risk factors

We identified modifiable risk factors for the distinct BMI trajectories (Figure 4 and eSupplement-Table 10). Maternal smoking during pregnancy (1.50 [0.88–2.54]) and absence of breastfeeding (1.56 [1.07–2.29]) was associated with increased odds of the *childhood onset obesity* trajectory. Maternal smoking during pregnancy made *persistent low BMI* less likely (0.64 [0.45–0.91]). Exposure to secondhand smoke showed increased odds of *early overweight resolving* (1.37 [0.98–1.91]). More physical activity between 6–18 years lowered the odds of *childhood onset obesity* (0.64 [0.44–0.93], ≥4 vs 0–3 hours/week) and *adolescent onset overweight* (0.75 [0.59–0.94]). Eating less fresh fruit between 4–18 years were weakly associated with higher odds of *adolescent onset overweight* (1.19 [0.92–1.54]).

Subgroup analysis by ethnic groups showed similar effect directions for these risk factors, but confidence intervals, mainly for South Asians, were wider (eSupplement-Tables 11 and 12).

## DISCUSSION

### Principal findings

In this large population-based cohort of children from Leicestershire we found five distinct trajectories of BMI development: *stable normal BMI* (47%); *persistent low BMI* (30%); *early overweight resolving* (8%)*; childhood onset obesity* (4%); and *adolescent onset overweight* (11%)*. Childhood onset obesity* started deviating from *stable normal BMI* at 2–4 years; *adolescent onset overweight* started deviating at 4–6 years. South Asian and White children showed the same five trajectories, but South Asians were more likely to follow *persistent low BMI*, *childhood onset obesity*, and *adolescent onset overweight* trajectories. Girls were at higher risk for these overweight and obesity trajectories. Children with less educated parents and those from socioeconomic deprived backgrounds were more likely to follow *childhood onset obesity*. Maternal smoking during pregnancy and no breastfeeding increased risk of *childhood onset obesity*. More physical activity (≥4 hours/week) protected against *childhood onset obesity* and *adolescent onset overweight*.

### Strengths and limitations

Main strengths of our study are the population-based study design, long follow-up, rich information on demographic, socioeconomic, perinatal, and lifestyle factors, and statistical methods taking advantage of the longitudinal design. We studied underserved populations by including children of South Asian ethnicity and from socioeconomically deprived backgrounds. We used IPW to reduce selection bias.

We were limited by partly using parent- and self-reports of BMI, mainly during adolescence, possibly underreporting high BMI values. This could have lowered the proportion of overweight and obesity trajectories and reduced our ability to observe the effects of exposures on these trajectories. We mitigated this by also using measured height and weight from routine healthcare. Because we did not assess adiposity status with additional measures (e.g., waist circumference or body fat percentage), ^32^ we may have misclassified participants. Misclassification may be differential because BMI underestimates body fat more strongly in South Asians than in Whites. ^33^ We mitigated this by also modelling BMI trajectories separately by ethnicity and sex. The number of BMI recordings varied, so participants with more BMI measurements contributed more strongly to the GBTM; this was mitigated by the fact that most children (78%) had 3 to 5 BMI measurements and contributed equally. And because we did not have information on parental adiposity status and genetic predispositions, the study may suffer from residual confounding. Compared to previous studies, we adjusted for a large set of exposures and hence reduced confounding. We included data from 1990–2010 and the prevalence of overweight and obesity in UK children has increased since. Hence, generalizability may be limited and the proportions of overweight and obesity trajectories might be higher in recent birth cohorts. Nevertheless, we think the described patterns of BMI development and effects of exposures remain valid.

### Comparison with other studies

Previous studies investigating BMI until adolescence or beyond described three to six trajectories including stable normal, above normal, persistent low, early increasing, late increasing, and accelerated resolving trajectories. ^7–9^ Early increasing trajectories started deviating from normal BMI at 2–6 years and late increasing ones between 6–14 years. We identified five similar trajectories in UK children that were comparable in Whites and South Asians, girls and boys. We narrowed the age of premature adiposity rebound in individuals with *childhood onset obesity* (2–4 years) and *adolescent onset overweight* (4–6 years). We also found that *early overweight resolving*, previously described as accelerated resolving, was related to perinatal parameters, in particular birth weight, and may resolve in adolescence.

Previous cross-sectional UK studies found obesity was more prevalent in South Asian ethnic minorities and socioeconomically deprived populations, ^1,2^ and two longitudinal studies between 7–17 years showed that socioeconomic inequalities widen in adolescence. ^3,4^ Cross-sectional prevalences however cannot directly be compared to longitudinal trajectories, but our results affirm that South Asians, girls, children with less educated parents, and those from socioeconomically deprived backgrounds more often follow overweight and obesity trajectories, marking them as at-risk for overweight and obesity. South Asian children have been reported to have higher cross-sectional underweight prevalences, ^1^ and our longitudinal findings align: South Asian children in our study were at higher risk for a *persistent low BMI* trajectory.

In previous UK studies, maternal smoking during pregnancy and no breastfeeding were identified as early-life risk factors for overweight and obesity in 3-year-olds and 11-year-olds. ^21,22^ We showed these remain risk factors for longitudinal trajectories of *childhood onset obesity*; this risk of obesity continues to 18 years. Maternal smoking during pregnancy reduces birth weight: but newborns rapidly catch-up and are at higher risk of overweight and obesity. ^23^ Rapid catch-up growth may explain why maternal smoking during pregnancy was associated with lower odds for the *persistent low BMI* trajectory in our study. Development of obesity may also be influenced by modifiable lifestyle factors, especially physical activity and diet. ^24,25^ We found that more physical activity protects against *childhood onset obesity* and *adolescent onset overweight*, though we could not exclude reverse causation since physical activity was reported on multiple questionnaires between 6–18 years. We found that lower fruit intake may raise the risk of *adolescent onset overweight*, but aggregated measures and limited power prevented us from evaluating diet in more detail; diet questions were only sent to subgroups. We did not study other potential risk factors including screen time, sedentary behaviour, sleep quality, and mental health. ^26–29^

### Implications

Public health policymakers need evidence to determine the best timing for interventions. We demonstrated that BMI increased at a higher rate during preschool (2–4 years) and early school age (4–6 years), opening windows and opportunities for action. UK obesity policies have been criticized for inadequately addressing social and ethnic aspects; ^30,31^ our findings underscore the need to incorporate ethnic and socioeconomic factors into future strategies, with a focus on reducing inequalities. In our study, South Asian children were at higher risk for obesity even after adjusting for demographic, socioeconomic, perinatal, and lifestyle risk factors, suggesting that additional determinants of obesity risk warrant further investigation of this disadvantaged population.

### Conclusions

In this large, longitudinal population-based, ethnically diverse cohort, we found that BMI in children with obesity starts to increase as early as 2–4 years, and at 4–6 years for adolescents with overweight. South Asian and socioeconomically deprived children were at higher risk of following overweight and obesity trajectories, indicating ethnic and social health inequalities beyond the factors usually considered. We suggest public health researchers and policymakers consider these inequalities when developing early-life obesity interventions.

## CONTRIBUTORS

CEK and Michael Silverman designed and conducted the Leicester Respiratory cohort studies. CEK and LL developed the idea for this analysis. LML prepared the data and performed the statistical analyses. LML and FNB accessed the data. CEK, FNB, BDS, and MG supervised the statistical analysis. All authors interpreted and discussed results. LML wrote the first draft of the manuscript, and all authors critically reviewed and revised it. All authors approved the final version of the manuscript. All authors had full access to the data in the study and were responsible for the decision to submit the manuscript. There were no funds or time allocated for patient and public involvement, so we could not involve participants or minoritised groups. We have invited participants to support the dissemination strategy of previous publications of the Leicester Respiratory Cohorts and do the same for this publication.

## Supporting information

STROBE checklist

GRoLTS checklist

## ACKNOWLEDGEMENTS

We would like to thank all families that participated in the Leicester Respiratory Cohorts. We would like to thank Professor Kali Tal for manuscript editing.

## COMPETING INTERESTS

CEK declares grants from the Swiss National Science Foundation for the submitted work; EAG reports grants from Wellcome Trust, NIHR Programme development grant, UKRI – SBRI, European Respiratory Society, University of Leicester, Midlands Asthma and Allergy Research Association, and Leicester, Leicestershire, Rutland ICB outside of the submitted work, payments or honoraria from Circassia, AstraZeneca, Sanofi, and Thorasys outside of the submitted work, and support for attending meetings from the European Respiratory Society. All authors have completed the ICMJE disclosure form and declare no other competing interests.

## DATA AVAILABILITY STATEMENT

Data may be made available to investigators upon request by email to the corresponding author, after approval of a proposal and with a signed data access agreement.

## FUNDING STATEMENT

Data collection and curation of the Leicester Respiratory Cohorts were supported by the Swiss National Science Foundation (grants: SNF PDFMP3-137033, 32003B-162820, 32003B-144068, 32003B-122341, PDFMP3-123162). The Leicester Respiratory Cohorts were also supported by Asthma UK, The Wellcome Trust, R&D Directorates of NHS Trusts, Children’s Research Fund, Medisearch (Leicester), and BUPA Foundation. LML was supported by a grant of the Swiss Personalized Health Network (NDS-2021-911 (SwissPedHealth)). The funder of the study had no role in study design, data collection, data analysis, data interpretation, or writing of the report.

## Notes

### Author Declarations

The Leicestershire Health Authority Research Ethics Committee approved the study (Ref numbers 4867, 5005, 04/Q2501/61, 07/H0407/70), and written consent was obtained from children and parents.

