## Supplementary Materials for "Ethnic and Social Health Inequalities in Body Mass Index Trajectories through Childhood and Adolescence: A Longitudinal Population-Based Study in Leicestershire UK"

### eSupplement Methods

#### Body mass index trajectory modelling

The four step approach of Ram and Grimm guided us through the body mass index (BMI) trajectory analysis: ^1^

i. **Problem definition:** We aimed to model BMI development trajectories from 0 to 18 years, either using Group-Based Trajectory Modelling (GBTM or Latent Class Growth Analysis [LCGA]), or to use Growth Mixture Modelling (GMM). GBTM models estimate class-specific mean trajectories, while GMM includes random effects to allow individual variation around the class mean. We chose GBTM because we were looking for distinct BMI development trajectories and sought to group participants accordingly. If we had used GMM with random effects, it might have returned fewer classes or less distinct trajectory patterns. After reviewing the literature, we chose to model raw BMI, which is a better measure of adiposity change and is easier to interpret than z-scores. ^2-4^ We did not include birth weight in our trajectory model because we did not have information on birth length and could not calculate BMI of newborns. We required participants to have at least three BMI values, one at 0–1 years and one at 3–10 years. Based on our review of the literature, we expected to find three to five distinct trajectories. ^5-7^
ii. **Model specification:** Previous studies mainly used polynomials (up to 3^rd^ degree) for age when modelling BMI trajectories; ^5^ Nummi et al. used splines for latent modelling of BMI trajectories with raw BMI. ^8^ We performed a grid search on a 1-class model to find a suitable function for estimating age patterns of BMI development in our study population. We compared models using up to 3^rd^ degree polynomials and splines with up to three knots, evaluating fit with Akaike information criterion (AIC), Bayesian information criterion (BIC), and judged biological plausibility by visually comparing to the UK-WHO growth charts. A 2^nd^ degree spline with two knots at 0.7 and 9.8 years provided the best fitting age pattern according to BIC, whereas AIC favoured a 2^nd^ degree spline function with three knots at 0.7, 10.3, and 13.8. Because both models showed good visual agreement with the UK-WHO growth charts, we chose the more parsimonious spline with two knots to specify GBTM models with 1 to 7 classes. To maintain model parsimony, we set the residual variance (variance-covariance matrix) to be equal across classes. We separately modelled girls and boys and White and South Asian ethnicities, then combined the whole study population in one model because the separate models showed similar trajectories (eSupplement-Figure 3).
iii. **Model estimation:** We estimated the models with the *lcmm* package in R, which employs full information maximum likelihood to determine model parameters. ^9^ We used 50 random starts and 20 iterations per random start for all models. We carried out the computations on UBELIX, the High Performance Computing (HPC) cluster at the University of Bern. ^10^
iv. **Model selection:** We evaluated model fit based on BIC and used likelihood ratio tests (LRT) and Lo-Mendell-Rubin LRTs to compare models. ^11^ We selected the optimal model based on model fit, class sizes, entropy levels, average posterior probabilities, proportion of participants with posterior probability ≥0.7, and biological plausibility. Model fit increased along with classes; the 7-class model fit best. We dropped the 7- and 6-class models because class sizes (1%) were small, entropy was less, and posterior probabilities were worse than for the 5-class model (Supplement-Figure 3 and Supplement-Table 6). The 5-class model fit better than the 4-class model, had lower BIC, significant Lo-Mendell-Rubin LRT, and also had better entropy, so the 5-class was the optimal model.

#### Identifying at-risk groups and risk factors: association of demographic, socioeconomic, perinatal, and lifestyle factors with BMI trajectories using multinomial logistic regression

We assessed the association of demographic, socioeconomic, perinatal, and lifestyle factors with BMI trajectories by using multinomial logistic regression to identify at-risk groups and risk factors. We used univariable multinomial logistic regression to estimate unadjusted odds ratios. To identify at-risk groups, we used multivariable multinomial logistic regression to estimate adjusted odds ratios of demographic, socioeconomic, and perinatal factors, mutually adjusting for all demographic, socioeconomic, and perinatal factors. To identify risk factors, we used multivariable multinomial logistic regression to estimate adjusted odds ratios of lifestyle factors, mutually adjusting for all demographic, socioeconomic, perinatal, and lifestyle factors.

##### Uncertainty of class membership

The GBTM model estimated posterior probabilities of every participant belonging to each BMI trajectory it identified. We took uncertainty of class membership into account when we estimated the multinomial logistic regression models. We weighted each participant’s contribution to the regression model by their posterior probability of membership for each of the identified BMI trajectories.

##### Reasoning for complete records analysis with inverse probability weighting

Some participants had no recorded measurements in the Leicestershire Health Authority Child Health Database, some questionnaires were sent only to subgroups, and some participants did not answer questionnaires completely or at all. Data for identifying at-risk groups and risk factors was missing in the outcome and in individual-level socioeconomic, perinatal, and lifestyle exposures. We evaluated several methods for handling missing data, using DAGs with missingness indicators (see eSupplement-Figure 1):

i. **Complete records analysis (CRA):** The analysis is conditioned on participants with available information on BMI trajectory. There is an open backwards path from BMI trajectory through missingness in BMI trajectory to demographic and socioeconomic factors, which leads to selection bias. Selection bias can be reduced by inverse probability weighting (IPW) on demographic and socioeconomic factors, removing the effects of demographic and socioeconomic factors on missingness in the BMI trajectory, closing the backwards path. Barlett et al. showed that exposure odds ratios estimated by logistic regression under CRA are asymptotically unbiased if the missingness of the outcome is only related to the outcome and the missingness of the exposures and confounders is only related to the exposures and confounders. ^12^ This was the case for our analysis, assuming that selection bias was kept to a minimum after IPW.
ii. **Multiple imputation (MI):** BMI trajectory likely affects its own missingness; demographic, socioeconomic, perinatal, and lifestyle factors likely affect their own missingness, e.g.: a participant with a high BMI may be less likely to report their BMI; a mother who smoked during pregnancy may be less likely to answer that question; a participant who is less physically active may be less likely to answer questions about physical activity. We concluded that the data were not missing at random (MNAR) and MI would lead to invalid estimates.

For these reasons, we used CRA with IPW but decided to not use MI to identify at-risk groups and risk factors for BMI trajectories with multinomial logistic regression.

#### Inverse probability weighting

To reduce selection bias, we used IPW on demographic and socioeconomic factors to weight participants included in the multinomial logistic regression analysis. We compared the demographic, socioeconomic, perinatal, and lifestyle characteristics of included and excluded participants (eSupplement-Table 2). We fitted a multivariable logistic regression model to estimate the probability of inclusion in the BMI trajectory analysis (vs exclusion) based on demographic and socioeconomic factors (eSupplement-Table 3), and used these probabilities to calculate stabilised IPW. We evaluated the IPW by calculating standardised differences by comparing included participants with all participants of the Leicester Respiratory Cohorts; we used the *cobalt* package in R. We used a love plot to compare standardised differences before and after the IPW (eSupplement-Figure 2). The standardised differences of demographic and socioeconomic characteristics were near zero (all <0.01); the differences were small for perinatal and lifestyle characteristics (all <0.1). We multiplied stabilised IPW with the posterior probabilities of class membership to account for uncertainty of class membership and used these total weights in our multinomial logistic regression analysis.

The Leicester Respiratory Cohorts sampled White children in 1990 and White and South Asian children 1998; in 1998, South Asian ethnicities were oversampled. We could not use weighting based on sampling rates to make our study population representative of the UK population because the Leicester Respiratory Cohorts did not sample children of other ethnicities and sampled South Asian children in 1998 but not in 1990.

#### eSupplement Figures and Tables

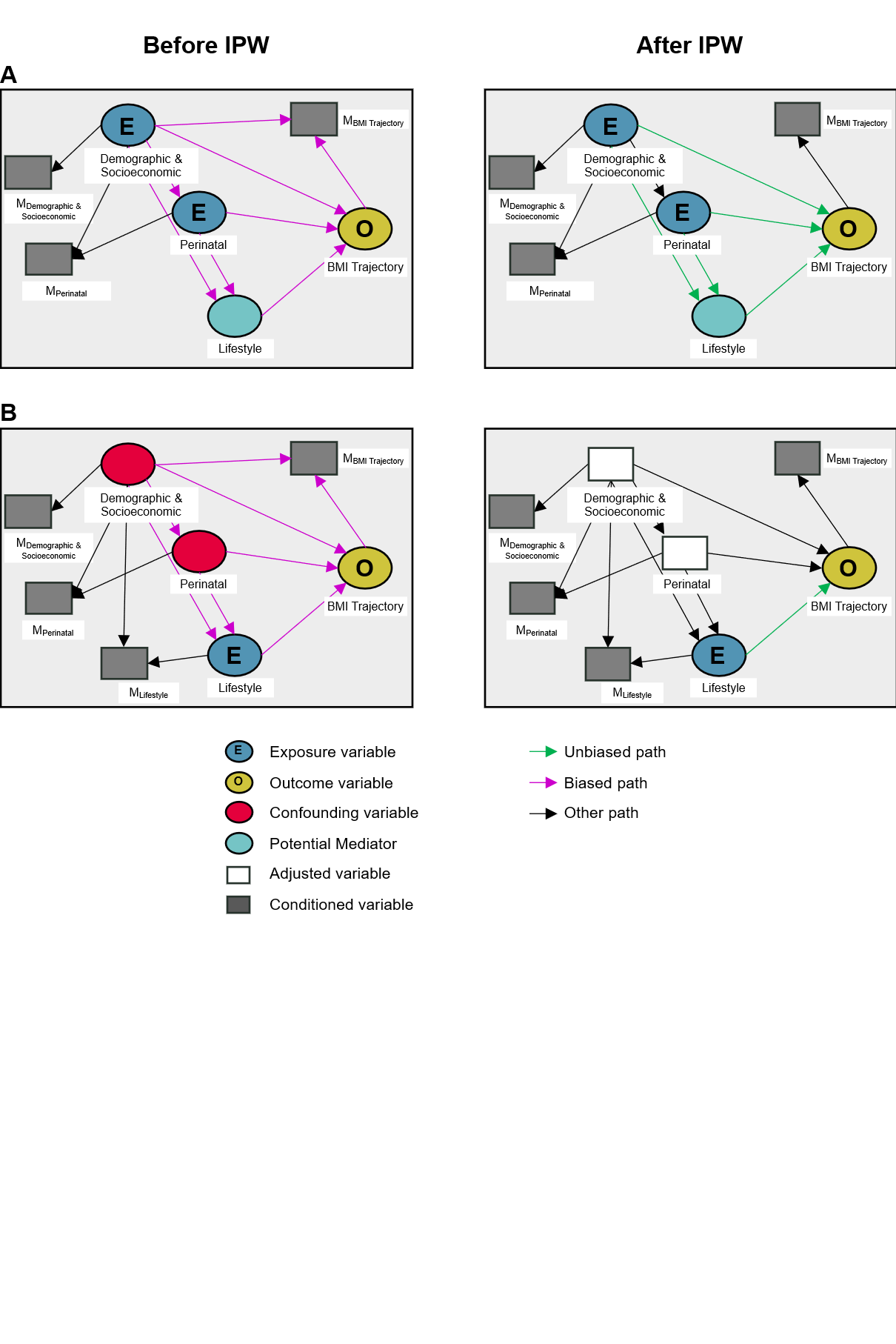

**eSupplement-Figure 1: Directed acyclic graphs (DAG) of A) identifying at-risk groups and B) identifying risk factors. Association of demographic, socioeconomic, perinatal, and lifestyle factors with BMI trajectories before and after inverse probability weighting.**
The analysis was conditioned on participants without missingness in BMI trajectory, which introduced potential selection bias through a backwards path to demographic and socioeconomic factors. After IPW, this backwards path was closed. Using multinomial logistic regression resulted in asymptotically unbiased estimates of adjusted odds ratios. ^12^ We did not draw potential residual confounding or reverse causation in this Figure (see the limitations section of the manuscript for further discussion).
Abbreviations: BMI—Body mass index; E—Exposure; IPW—Inverse probability weighting; O—Outcome.

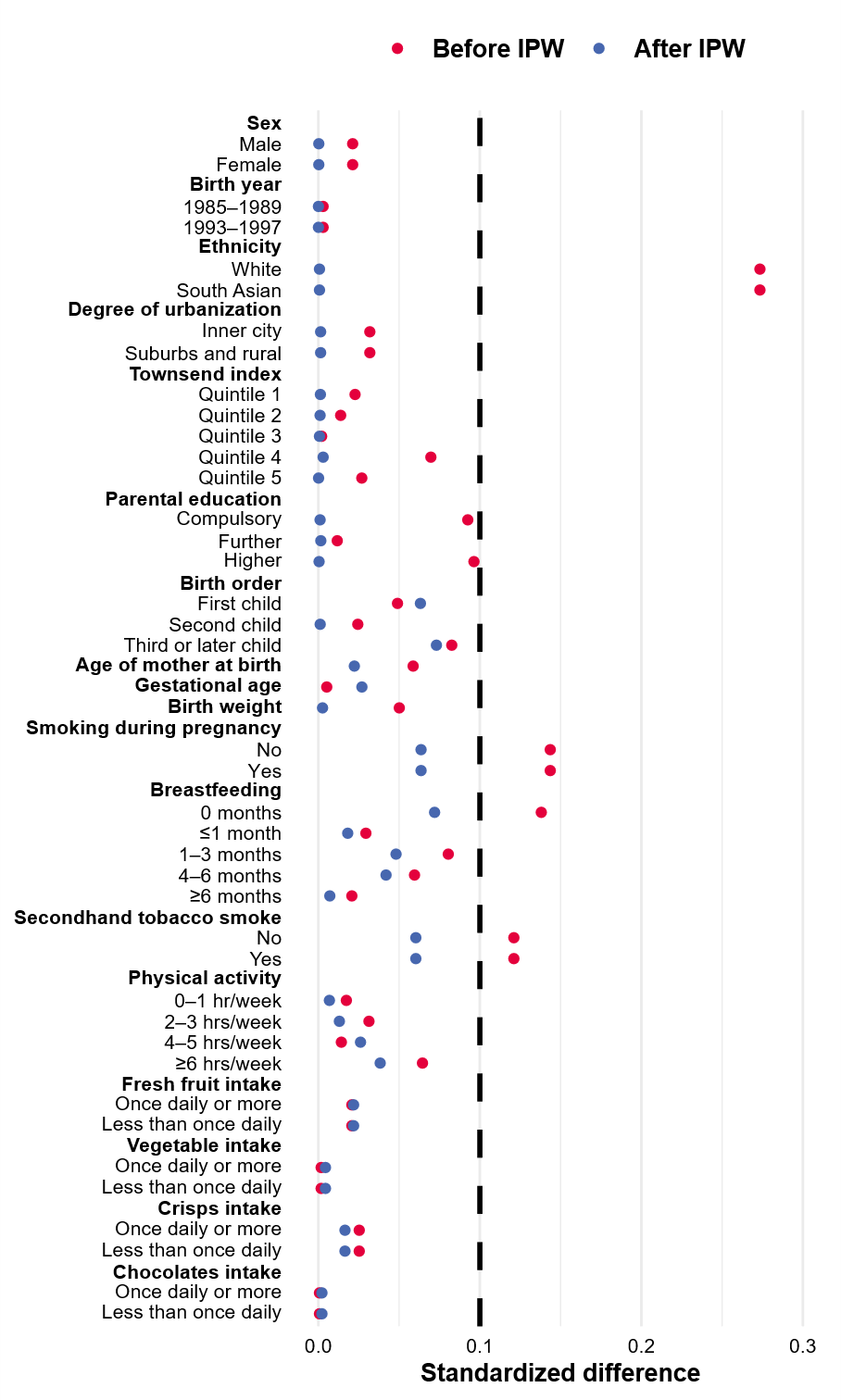

**eSupplement-Figure 2: Standardised differences when participants included in the BMI trajectory analysis were compared to all participants of the Leicester Respiratory Cohorts, before and after inverse probability weighting.**
Standardized differences for this plot were calculated with the *cobalt* package in R.
We used inverse probability weighting to reduce selection bias of participants included in the BMI trajectory analysis (see eSupplement Methods.
Abbreviations: IPW—inverse probability weighting.

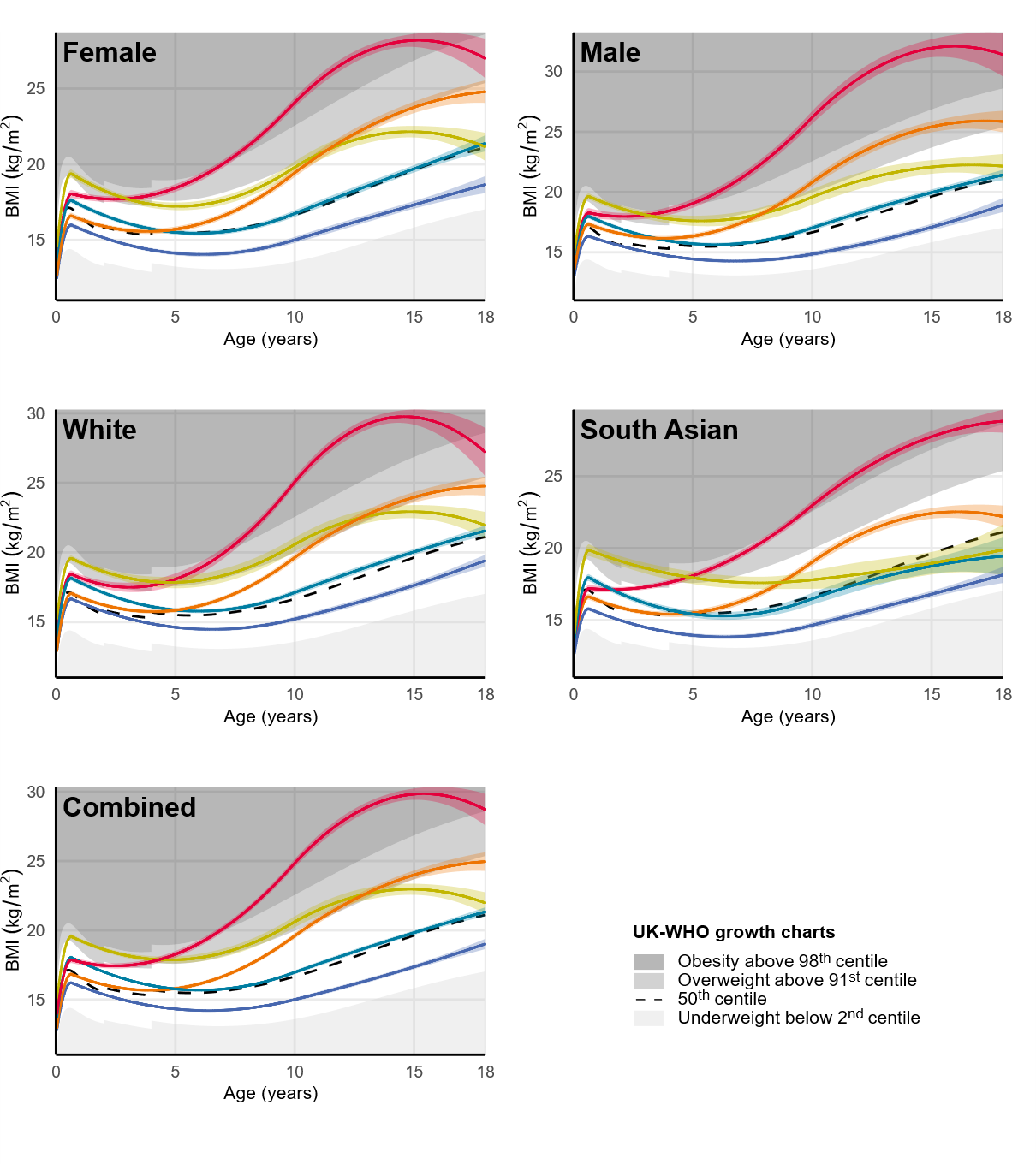

**eSupplement-Figure 3: Group-based BMI trajectory models estimated separately for girls, boys, White, and South Asian participants of the Leicester Respiratory Cohorts.**
The plotted trajectories are predicted marginal means and 95% confidence intervals of the Group-Based Trajectory Models. The plotted centiles in the “White”, “South Asian”, and “Combined” panels are averages of the girl and boy centiles of the UK-WHO growth charts. We combined all participants for the final model because the separate trajectories in girls and boys and in Whites and South Asians were visually comparable.
Abbreviations: BMI—Body mass index

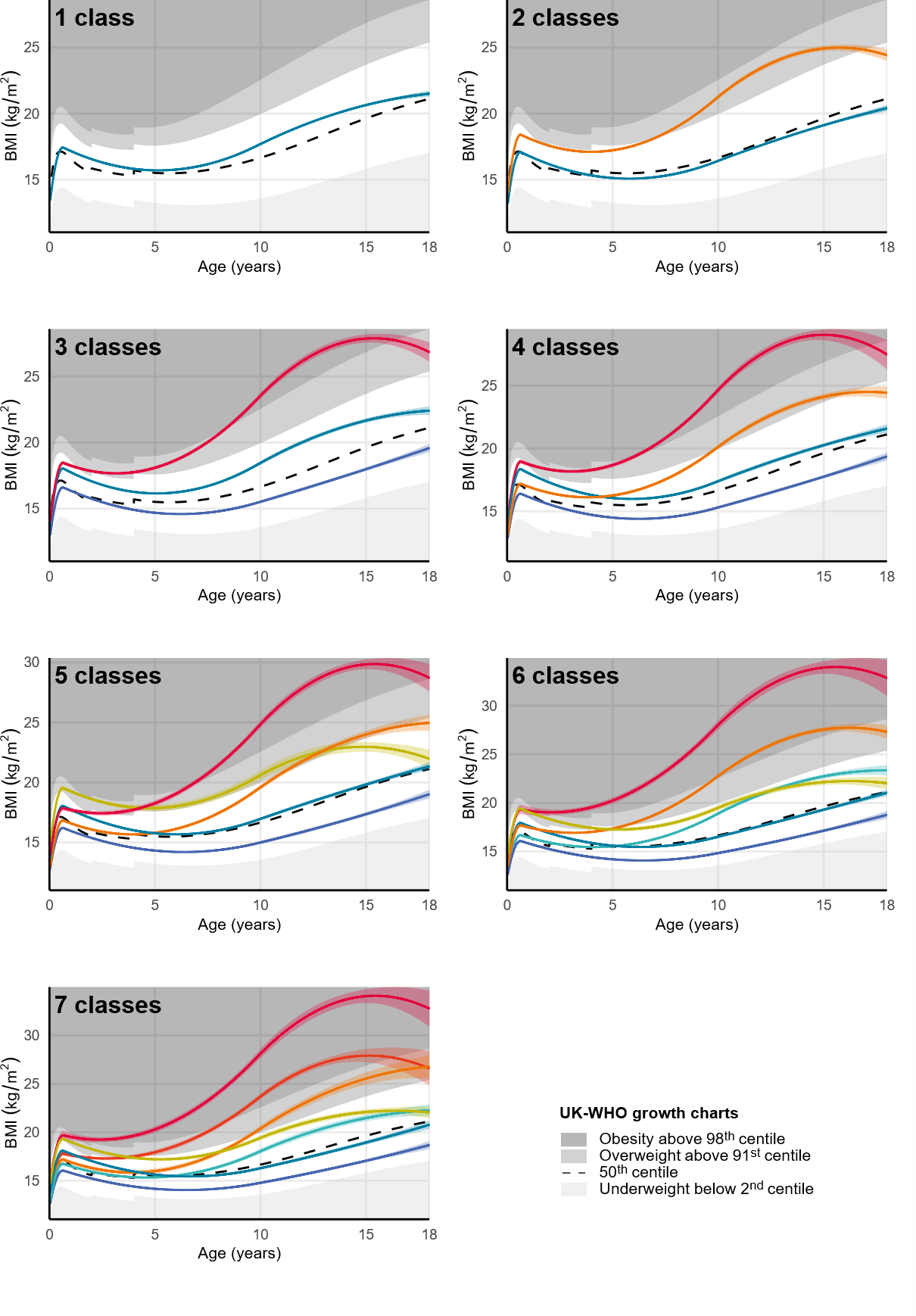

**eSupplement-Figure 4: Group-based BMI trajectory models from one to seven classes in participants of the Leicester Respiratory Cohorts.**
The plotted trajectories are predicted marginal means and 95% confidence intervals of the Group-Based Trajectory Models. The plotted centiles are averages of the girl and boy centiles of the UK-WHO growth charts.
Abbreviations: BMI—body mass index

**eSupplement-Table 1: Summary of questionnaires, clinical study visits, and linked routine data in the Leicester Respiratory Cohorts.** Adapted from Kuehni et al. ^13^

| **Year** | **Participants addressed** | **Age of participants (years) by cohort** | | |
| --- | --- | --- | --- | --- |
|  |  | 1990 | 1998(a) | 1998(b) |
| **Questionnaires** | | | | |
| 1990 | All families of 1990 cohort. | 0–4 |  |  |
| 1992-1994 | Nested sample of 1990 cohort with wheeze or cough in 1990 (n=448).  Nested sample of asymptomatic 1990 cohort (n=347). | 2–8 |  |  |
| 1998 | All families of 1990 and 1998 cohorts. | 8–12 | 1–4 | 1 |
| 1999 | All families of 1998(b) cohort with responses in 1998. |  |  | 2 |
| 2001 | All families of 1998 cohorts |  | 4–7 | 4 |
| 2003 | All families of 1990 and 1998 cohorts. Children/adolescents of 1990 cohort answered questionnaires themselves. | 13–17 | 6–9 | 6 |
| 2005-2007 | All families of 1998 cohorts. Both parents and children answered separate questionnaires. |  | 8–13 | 8–9 |
| 2010 | All families of 1990 and 1998 cohorts.  Adolescents of all cohorts answered questionnaires themselves. | 20–24 | 13–16 | 13 |
| 2012 | All families of 1990 and 1998 cohorts.  Adolescents of all cohorts answered questionnaires themselves. | 22–26 | 15–18 | 15 |
| **Clinical study visits** | | | | |
| 1992-1994 | Nested sample of 1990 cohort with wheeze or cough in 1990 (n=448).  Nested sample of asymptomatic 1990 cohort (n=347). | 2–8 |  |  |
| 2005-2007 | All families of 1998 cohorts with responses in 1998 and one additional response. |  | 8–13 | 8 |
| **Routine health care data** | | | | |
| 1990–2006 | All children: routine health care data from the Leicestershire Health Authority Child Health Database:   - Demographic information - Perinatal information - Anthropometric measurements from preventive well-child visits (birth, 1–2 weeks, 6–8 weeks, 9–12 months, 2–2.5 years), and other health related events during childhood. | 0–15 | 0–11 | 0–8 |
| 1990 and 1998 | All families: data from UK census 2001 on most detailed geographic level (output area) linked to the postal code of the participants at beginning of the study.^1^   - Degree of urbanisation - Townsend index of socioeconomic deprivation | 0–4 | 1–4 | 1 |

^1^ Linked Townsend index of socioeconomic deprivation: We downloaded the Townsend index calculated from 2001 UK censuses from UK Data Service Open. ^14^ We linked this index to the participants’ postal code at baseline. The Townsend index was not available for output areas from the census 1991, so we also linked the 2001 Townsend index to the baseline postal code of the 1990 cohort.

**eSupplement-Table 2: Questions from the Leicester Respiratory cohorts and coding used in this analysis**

| **Variable** | **Question** | **Variable coding** |
| --- | --- | --- |
| Parental education | Father or male guardian: At what age did he finish his full-time education? _________ (years)  Mother or female guardian At what age die she finish her full-time education? _________ (years) | Compulsory education (≤16 years)  Further education (17–19 years)  Higher education (≥20 years)  Based on the higher value from either parent. |
| Smoking during pregnancy | Mother or female guardian  Did she smoke cigarettes during the year in which your child was born?   - Yes - No | No  Yes |
| Breastfeeding | Was your child breastfed as a baby?   - Yes - No   If yes, for how long?  _________ (months) | No  ≤1 month  1–3 months  4–6 months  ≥6 months  For regression models we avoided small categories and coded it as:  Yes  No |
| Secondhand tobacco smoke | Does the child’s mother smoke cigarettes?   - Yes - No   Does the child’s father/Do any other household members smoke cigarettes?   - Yes - No | No  Yes  For participants with answers in multiple questionnaires we coded “No, never” or “Yes, ever” |
| Vigorous physical activity | On average, how many hours a week does your child/do you spend on sports, games or vigorous physical activity? *(vigorous activities cause a large increase in breathing and heart rate; they may include heavy lifting, heavy yard work, digging, aerobics, fast cycling, or running)*   - 0 to 1 hr - 2 to 3 hrs - 4 to 5 hrs - 6 hrs or more | 0–1 hours per week  2–3 hours per week  4–5 hours per week  ≥6 hours per week  Question was introduced in 2003. For participants in Cohort 1 (1990) we took the answer from 2003 (ages 13–17 years).  For participants in Cohort 2 (1998a) and Cohort 3 (1998b) we calculated an average of the participant’s answers from 2003 to 2012 (ages 6–18 years).  For regression models we avoided small categories and coded it as:  0–3 hours per week  ≥4 hours per week |
| Fresh fruit intake | In the last 12 months, how often did your child/did you on average eat fresh fruit?   - Twice daily - Daily - 2 to 4 times a week - Less than twice a week | Less than once daily  Once daily or more  Question was introduced in 2001. For participants in Cohort 1 (1990) we took the answer from 2003 (ages 13–17 years).  For participants in Cohort 2 (1998a) and Cohort 3 (1998b) we calculated an average of the participant’s answers from 2001 to 2012 (ages 4–18 years). |
| Vegetable intake | In the last 12 months, how often did your child/did you on average eat vegetables (not potatoes)?   - Twice daily - Daily - 2 to 4 times a week - Less than twice a week | Less than once daily  Once daily or more  Question was introduced in 2001. For participants in Cohort 1 (1990) we took the answer from 2003 (ages 13–17 years).  For participants in Cohort 2 (1998a) and Cohort 3 (1998b) we calculated an average of the participant’s answers from 2001 to 2012 (ages 4–18 years). |
| Crisps intake | In the last 12 months, how often did your child/did you on average eat crisps or other salty snacks?   - Twice daily - Daily - 2 to 4 times a week - Less than twice a week | Less than once daily  Once daily or more  Question was introduced in 2001. For participants in Cohort 1 (1990) we took the answer from 2003 (ages 13–17 years).  For participants in Cohort 2 (1998a) and Cohort 3 (1998b) we calculated an average of the participant’s answers from 2001 to 2012 (ages 4–18 years). |
| Chocolates intake | In the last 12 months, how often did your child/did you on average eat chocolates or sweets?   - Twice daily - Daily - 2 to 4 times a week - Less than twice a week | Less than once daily  Once daily or more  Question was introduced in 2001. For participants in Cohort 1 (1990) we took the answer from 2003 (ages 13–17 years).  For participants in Cohort 2 (1998a) and Cohort 3 (1998b) we calculated an average of the participant’s answers from 2001 to 2012 (ages 4–18 years). |

**eSupplement-Table 3: Characteristics of the participants of the Leicester Respiratory Cohorts in total, included vs excluded in the BMI trajectory analysis.**

|  | | **Total,** n (%) 10,350 (100) | **Included**, n (%)  5,571 (54) | **Excluded,** n (%) 4,779 (46) | **P Value^1^** |
| --- | --- | --- | --- | --- | --- |
| **DEMOGRAPHIC AND SOCIOECONOMIC** | | | | | |
| **Sex**, n (%) | Male | 5,355 (52) | 2,910 (52) | 2,445 (51) | 0.2808 |
|  | Female | 4,994 (48) | 2,661 (48) | 2,333 (49) |  |
|  | *Missing* | *1 (<0.1)* | *-* | *1 (<0.1)* |  |
| **Birth year**, n (%) | 1985–1989 | 1,650 (16) | 891 (16) | 759 (16) | 0.8772 |
|  | 1993–1997 | 8,700 (84) | 4,680 (84) | 4,020 (84) |  |
| **Ethnicity**, n (%) | White | 7,750 (75) | 3,871 (69) | 3,879 (81) | <0.0001 |
|  | South Asian | 2,600 (25) | 1,700 (31) | 900 (19) |  |
| **Parental education^2^**, n (%) | Compulsory education | 3,472 (44) | 1,849 (42) | 1,623 (46) | <0.0001 |
|  | Further education | 2,654 (33) | 1,493 (34) | 1,161 (33) |  |
|  | Higher education | 1,828 (23) | 1,100 (25) | 728 (21) |  |
|  | *Missing* | *2,396 (23)* | *1,129 (20)* | *1,267 (27)* |  |
| **Degree of urbanisation^3^**, n (%) | Inner city | 5,389 (52) | 2,860 (51) | 2,529 (53) | 0.2083 |
|  | Suburbs and rural | 4,961 (48) | 2,711 (49) | 2,250 (47) |  |
| **Townsend index^4^** | Median [IQR] | -0.5 (-2.6, 2.9) | -0.4 (-2.5, 2.7) | -0.6 (-2.6, 3.0) | 0.2469 |
|  | Quintile, n (%) |  |  |  |  |
|  | 1 (low deprivation) | 2,044 (20) | 1,077 (19) | 967 (20) | 0.0110 |
|  | 2 | 2,041 (20) | 1,084 (19) | 957 (20) |  |
|  | 3 | 1,854 (18) | 996 (18) | 858 (18) |  |
|  | 4 | 1,900 (18) | 1,092 (20) | 808 (17) |  |
|  | 5 (high deprivation) | *2,511 (24)* | *1,322 (24)* | *1,189 (25)* |  |
| **PERINATAL** | | | | | |
| **Birth order**, n (%) | First child | 3,636 (41) | 2,001 (42) | 1,635 (39) | 0.0004 |
|  | Second child | 3,079 (35) | 1,675 (35) | 1,404 (34) |  |
|  | Third or later child | 2,209 (25) | 1,104 (23) | 1,105 (27) |  |
|  | *Missing* | *1,426 (14)* | *791 (14)* | *635 (13)* |  |
| **Age of mother at birth (years)** | Mean (SD) | 28.2 (5.3) | 28.3 (5.2) | 28.0 (5.4) | 0.0051 |
|  | *Missing* | *1,168 (11)* | *614 (11)* | *554 (12)* |  |
| **Gestational age (weeks)** | Mean (SD) | 39.2 (1.9) | 39.2 (1.9) | 39.2 (2.0) | 0.8059 |
|  | *Missing* | *1,427 (14)* | *787 (14)* | *640 (13)* |  |
| **Birth weight (grams)** | Mean (SD) | 3,295 (575) | 3,282 (565) | 3,311 (586) | 0.0121 |
|  | *Missing* | *251 (2)* | *165 (3)* | *86 (2)* |  |
| **LIFESTYLE** | | | | | |
| **Smoking during pregnancy**, n (%) | No | 6,515 (82) | 3,749 (84) | 2,766 (78) | <0.0001 |
|  | Yes | 1,468 (18) | 710 (16) | 758 (22) |  |
|  | *Missing* | *2,367 (23)* | *1,112 (20)* | *1,255 (26)* |  |
| **Breastfeeding**, n (%) | No | 2,914 (40) | 1,527 (38) | 1,387 (44) | <0.0001 |
|  | ≤1 month | 794 (11) | 465 (11) | 329 (11) |  |
|  | 1–3 months | 1,341 (19) | 813 (20) | 528 (17) |  |
|  | 4–6 months | 884 (12) | 534 (13) | 350 (11) |  |
|  | ≥6 months | 1,267 (18) | 730 (18) | 537 (17) |  |
|  | *Missing* | *3,150 (30)* | *1,502 (27)* | *1,648 (34)* |  |
| **Secondhand tobacco smoke^5^**, n (%) | No | 5,045 (55) | 2,940 (58) | 2,105 (52) | <0.0001 |
|  | Yes | 4,153 (45) | 2,171 (42) | 1,982 (48) |  |
|  | *Missing* | *1,152 (11)* | *460 (8.3)* | *692 (14)* |  |
| **Vigorous physical activity^6^**, n (%) | 0–1 hours per week | 735 (12) | 451 (12) | 284 (11) | 0.0893 |
|  | 2–3 hours per week | 2,211 (35) | 1,354 (36) | 857 (34) |  |
|  | 4–5 hours per week | 1,958 (31) | 1,189 (31) | 769 (31) |  |
|  | ≥6 hours per week | 1,430 (23) | 820 (21) | 610 (24) |  |
|  | *Missing* | *4,016 (39)* | *1,757 (32)* | *2,259 (47)* |  |
| **Fresh fruit intake^7^**, n (%) | Once daily or more | 4,913 (67) | 2,867 (67) | 2,046 (67) | 0.3816 |
|  | Less than once daily | 2,430 (33) | 1,444 (33) | 986 (33) |  |
|  | *Missing* | *3,007 (29)* | *1,260 (23)* | *1,747 (37)* |  |
| **Vegetable intake^7^**, n (%) | Once daily or more | 1,686 (64) | 1,061 (64) | 625 (64) | 0.9655 |
|  | Less than once daily | 942 (36) | 592 (36) | 350 (36) |  |
|  | *Missing* | *7,722 (75)* | *3,918 (70)* | *3,804 (80)* |  |
| **Crisps intake^7^**, n (%) | Once daily or more | 2,145 (49) | 1,298 (49) | 847 (50) | 0.4156 |
|  | Less than once daily | 2,202 (51) | 1,359 (51) | 843 (50) |  |
|  | *Missing* | *6,003 (58)* | *2,914 (52)* | *3,089 (65)* |  |
| **Chocolates intake^7^**, n (%) | Once daily or more | 1,914 (44) | 1,169 (44) | 745 (44) | 0.9813 |
|  | Less than once daily | 2,430 (56) | 1,485 (56) | 945 (56) |  |
|  | *Missing* | *6,006 (58)* | *2,917 (52)* | *3,089 (65)* |  |

^1^P Value: We used Pearson’s chi-squared tests for categorical, t-test for normally distributed, and Kruskal-Wallis tests for non-normally distributed variables. ^2^Parental education based on the age at completion of full-time education, taking the higher value of either parent. Compulsory education is ≤16 years of age; further education is 17–19 years; higher education is ≥20 years. ^3^Degree of urbanisation: inner city includes postcodes LE1–LE5. ^4^Townsend index of socioeconomic deprivation for the year 2001. ^5^Secondhand tobacco smoke exposure between 0–18 years due to smoking of household member. ^6^Vigorous physical activity: average of a participant’s answers at ages 6–18 years. ^7^Dietary intake: average of a participant’s answers at ages 4–18 years. See eSupplement Materials for coding of questionnaire variables. Abbreviations: BMI—Body mass index; CI—confidence interval; IQR—inter-quartile range; SD—standard deviation.

**eSupplement-Table 4: Logistic regression model for inverse probability weighting of participants of the Leicester Respiratory Cohorts included in the BMI analysis.**

| **Demographic and socioeconomic characteristic** | **aOR [95% CI]** | **Std. error** | **P Value** |
| --- | --- | --- | --- |
| **Sex** |  |  |  |
| Male | ref |  |  |
| Female | 0.95 [0.88–1.03] | 1.04 | 0.1908 |
| **Birth year** |  |  |  |
| 1993–1997 | ref |  |  |
| 1985–1989 | 1.19 [1.07–1.33] | 1.06 | 0.0020 |
| **Ethnicity** |  |  |  |
| White | ref |  |  |
| South Asian | 2.48 [2.22–2.77] | 1.06 | <0.0001 |
| **Parental education^1^** |  |  |  |
| Compulsory education | ref |  |  |
| Further education | 1.04 [0.93–1.15] | 1.05 | 0.5215 |
| Higher education | 1.11 [0.98–1.25] | 1.06 | 0.1011 |
| Missing | 0.66 [0.59–0.73] | 1.06 | <0.0001 |
| **Degree of urbanisation** |  |  |  |
| Inner city | ref |  |  |
| Suburbs and rural | 1.46 [1.32–1.61] | 1.05 | <0.0001 |
| **Townsend index quintile^2^** |  |  |  |
| 1 (low deprivation) | ref |  |  |
| 2 | 1.04 [0.92–1.18] | 1.07 | 0.5566 |
| 3 | 1.07 [0.94–1.22] | 1.07 | 0.2853 |
| 4 | 1.25 [1.09–1.44] | 1.07 | 0.0016 |
| 5 (high deprivation) | 1.07 [0.93–1.23] | 1.07 | 0.3542 |

We fitted a multivariable logistic regression model to estimate the probability of inclusion in the BMI trajectory analysis (vs exclusion) based on demographic and socioeconomic factors (eSupplement-Table 3), and used these probabilities to calculate stabilised IPW. One participant not included in the BMI trajectory analysis was excluded from the IPW model because of missing information on sex.
Abbreviations: aOR—adjusted odds ratio; CI—confidence interval.

**eSupplement-Table 5: Number of BMI measurements per participant of the Leicester Respiratory Cohorts included in the BMI trajectory analysis.**

|  | **Participants,** n (%)  5,571 (100) |
| --- | --- |
| **Number of BMI measurements**, n (%) |  |
| 3 | 1,389 (25) |
| 4 | 1,769 (32) |
| 5 | 1,213 (22) |
| 6 | 709 (13) |
| 7 | 354 (6) |
| 8 | 109 (2) |
| 9 | 19 (<1) |
| 10 | 6 (<1) |
| 11 | 2 (<1) |
| 12 | 1 (<1) |
| Median [IQR] | 4 [4, 5] |

Abbreviations: BMI—Body mass index; IQR—Inter-quartile range

**eSupplement-Table 6: Number of BMI measurements by age year of participants in the Leicester Respiratory Cohorts included in the BMI trajectory analysis.**

|  | **Total number of BMI measurements,** n (%)  25,177 (100) |
| --- | --- |
| **Age (years)**, n (%) |  |
| 0 | 8,060 (32) |
| 1 | 3,488 (14) |
| 2 | 623 (2) |
| 3 | 4,848 (19) |
| 4 | 299 (1) |
| 5 | 1,686 (7) |
| 6 | 1,367 (5) |
| 7 | 387 (2) |
| 8 | 271 (1) |
| 9 | 843 (3) |
| 10 | 499 (2) |
| 11 | 267 (1) |
| 12 | 361 (1) |
| 13 | 656 (3) |
| 14 | 712 (3) |
| 15 | 316 (1) |
| 16 | 315 (1) |
| 17 | 188 (<1) |

Abbreviations: BMI—Body mass index

**eSupplement-Table 7: Indices of model fit for the Group-Based Trajectory Models of BMI development trajectories in the Leicester Respiratory Cohorts.**

| **# classes** | **# parameters** | **BIC** | **Entropy** | **Class percentages** | **Average posterior probability** | **Proportion of participants with posterior probability ≥ 0.7** |
| --- | --- | --- | --- | --- | --- | --- |
| 1 | 6 | 108351 | 1.000 | 100 | 1 | 1 |
| 2 | 12 | 102515 | 0.690 | 75, 25 | 0.92, 0.85 | 0.91, 0.78 |
| 3 | 18 | 100358 | 0.655 | 46, 45, 8 | 0.80, 0.86, 0.88 | 0.69, 0.80, 0.81 |
| 4 | 24 | 99395 | 0.631 | 41, 40, 13, 6 | 0.79, 0.81, 0.87, 0.75 | 0.66, 0.71, 0.81, 0.59 |
| 5 | 30 | 98786 | 0.641 | 47, 30, 11, 8, 4 | 0.74, 0.78, 0.67, 0.79, 0.85 | 0.58, 0.66, 0.62, 0.67, 0.76 |
| 6 | 36 | 98138 | 0.638 | 42, 24, 14, 12, 6, 1 | 0.70, 0.73, 0.71, 0.87, 0.84, 0.76 | 0.48, 0.53, 0.51, 0.79, 0.76, 0.62 |
| 7 | 42 | 97948 | 0.611 | 35, 22, 19, 13, 5, 4, 1 | 0.72, 0.60, 0.66, 0.80, 0.74, 0.86, 0.76 | 0.52, 0.34, 0.39, 0.67, 0.58, 0.76, 0.62 |

Abbreviations: BIC—Bayesian information criterion; BMI—Body mass index

**eSupplement-Table 8: Unadjusted associations of demographic, socioeconomic, perinatal, and lifestyle factors with BMI trajectories in participants of the Leicester Respiratory Cohorts.**

|  | **Univariable models with outcome BMI trajectory**, OR [95% CI] | | | | |
| --- | --- | --- | --- | --- | --- |
|  | Stable normal BMI, n (%) 2,625 (47) | Persistent low BMI, n (%) 1,648 (30) | Early overweight resolving, n (%) 442 (8) | Childhood onset obesity, n (%) 235 (4) | Adolescent onset overweight, n (%) 621 (11) |
| **DEMOGRAPHIC AND SOCIOECONOMIC** | | | | | |
| **Sex**, ref: Male |  |  |  |  |  |
| Female | ref | 1.53 [1.34–1.74] | 0.96 [0.79–1.16] | 1.26 [0.99–1.60] | 1.43 [1.23–1.66] |
| **Birth year**, ref: 1993–1997 |  |  |  |  |  |
| 1985–1989 | ref | 0.77 [0.64–0.92] | 1.15 [0.90–1.46] | 0.64 [0.45–0.92] | 0.76 [0.61–0.94] |
| **Ethnicity**, ref: White |  |  |  |  |  |
| South Asian | ref | 2.42 [2.08–2.80] | 0.75 [0.58–0.98] | 1.52 [1.15–2.00] | 1.62 [1.36–1.94] |
| **Parental education^1^**, ref: Compulsory education |  |  |  |  |  |
| Further education | ref | 1.11 [0.93–1.31] | 0.90 [0.70–1.15] | 0.97 [0.71–1.31] | 1.01 [0.82–1.23] |
| Higher education | ref | 1.23 [1.01–1.48] | 0.90 [0.68–1.20] | 0.71 [0.49–1.04] | 1.09 [0.87–1.36] |
| **Degree of urbanisation^2^**, ref: Inner city |  |  |  |  |  |
| Suburbs and rural | ref | 0.63 [0.55–0.72] | 1.01 [0.83–1.22] | 0.72 [0.57–0.91] | 0.72 [0.62–0.84] |
| **Townsend quintile^3^**, ref: 1 (low deprivation) |  |  |  |  |  |
| 2 | ref | 1.05 [0.85–1.30] | 0.92 [0.69–1.23] | 1.35 [0.90–2.02] | 1.08 [0.85–1.38] |
| 3 | ref | 1.22 [0.99–1.52] | 1.15 [0.86–1.53] | 1.45 [0.96–2.19] | 1.10 [0.86–1.42] |
| 4 | ref | 1.55 [1.25–1.91] | 0.83 [0.61–1.14] | 1.76 [1.18–2.63] | 1.31 [1.02–1.68] |
| 5 (high deprivation) | ref | 1.54 [1.27–1.88] | 0.90 [0.67–1.20] | 1.80 [1.23–2.63] | 1.44 [1.14–1.81] |
| **PERINATAL** | | | | | |
| **Birth order**, ref: First child |  |  |  |  |  |
| Second child | ref | 0.92 [0.79–1.08] | 0.92 [0.72–1.17] | 1.20 [0.90–1.60] | 0.90 [0.75–1.09] |
| Third or later child | ref | 0.95 [0.79–1.13] | 1.07 [0.82–1.39] | 1.13 [0.81–1.57] | 0.92 [0.74–1.13] |
| **Age of mother at birth (years)** |  |  |  |  |  |
| Per 1 SD increase: 5.3 years | ref | 0.96 [0.90–1.03] | 1.01 [0.91–1.12] | 1.00 [0.88–1.13] | 0.96 [0.89–1.04] |
| **Gestational age (weeks)** |  |  |  |  |  |
| Per 1 SD increase: 1.9 weeks | ref | 0.92 [0.86–0.99] | 1.18 [1.05–1.33] | 1.01 [0.88–1.15] | 0.96 [0.88–1.04] |
| **Birth weight (grams)** |  |  |  |  |  |
| Per 1 SD increase: 565 grams | ref | 0.65 [0.61–0.70] | 1.47 [1.32–1.63] | 1.06 [0.93–1.20] | 0.79 [0.73–0.86] |
| **LIFESTYLE** | | | | | |
| **Smoking during pregnancy**, ref: No |  |  |  |  |  |
| Yes | ref | 0.64 [0.51–0.79] | 1.28 [0.98–1.66] | 1.62 [1.18–2.22] | 1.01 [0.81–1.27] |
| **Breastfeeding**, ref: Yes |  |  |  |  |  |
| No | ref | 1.06 [0.91–1.25] | 0.97 [0.77–1.22] | 0.69 [0.52–0.92] | 0.96 [0.80–1.15] |
| **Secondhand tobacco smoke^4^**, ref: No |  |  |  |  |  |
| Yes | ref | 0.83 [0.72–0.95] | 1.34 [1.10–1.63] | 1.37 [1.07–1.76] | 1.10 [0.94–1.30] |
| **Vigorous physical activity^5^**, ref: 0–3 hours per week |  |  |  |  |  |
| ≥4 hours per week | ref | 0.8 [0.68–0.94] | 0.97 [0.76–1.22] | 0.64 [0.48–0.86] | 0.70 [0.58–0.84] |
| **Fresh fruits intake^6^**, ref: Once daily or more |  |  |  |  |  |
| Less than once daily | ref | 1.02 [0.88–1.20] | 1.18 [0.94–1.48] | 0.79 [0.59–1.07] | 1.07 [0.89–1.29] |
| **Vegetable intake^6^**, ref: Once daily or more |  |  |  |  |  |
| Less than once daily | ref | 1.21 [0.94–1.55] | 1.03 [0.71–1.50] | 1.31 [0.79–2.18] | 1.10 [0.82–1.46] |
| **Crisps intake^6^**, ref: Once daily or more |  |  |  |  |  |
| Less than once daily | ref | 0.90 [0.75–1.09] | 1.03 [0.78–1.38] | 0.91 [0.64–1.28] | 0.87 [0.70–1.08] |
| **Chocolates intake^6^**, ref: Once daily or more |  |  |  |  |  |
| Less than once daily | ref | 0.89 [0.74–1.08] | 1.14 [0.86–1.53] | 1.02 [0.72–1.45] | 1.08 [0.86–1.34] |

Presented are odds ratios with 95% confidence intervals from univariable multinomial logistic regression. The number of participants for each exposure depends on the missingness in the exposure, see Table 1 and Table 2. We recoded breastfeeding and physical activity to avoid small categories. We took the uncertainty of class membership into account by weighting each participant’s contribution to the regression model by their posterior probability of membership for each of the five BMI trajectories. We used inverse probability weighting to reduce selection bias arising from differences between participants included in the BMI trajectory analysis (vs excluded), see eSupplement Methods.
^1^Parental education based on age at completion of full-time education, taking the higher value of either parent. Compulsory education is ≤16 years of age; further education is 17–19 years; higher education is ≥20 years. ^2^Degree of urbanisation: inner city includes postcodes LE1–LE5. ^3^Townsend index of socioeconomic deprivation for the year 2001. ^4^Secondhand tobacco smoke exposure between 0–18 years due to smoking of household member. ^5^Vigorous physical activity: average of a participant’s answers at ages 6–18 years. ^6^Dietary intake: average of a participant’s answers at ages 4–18 years. See eSupplement-Table 2 for coding of questionnaire variables.
Abbreviations: OR—odds ratio; BMI—body mass index; CI—confidence interval; SD—standard deviation.

**eSupplement-Table 9: Identification of at-risk groups: Adjusted associations of demographic, socioeconomic, and perinatal factors with BMI trajectories in participants of the Leicester Respiratory Cohorts.**

|  | **Multivariable model with outcome BMI trajectory**, aOR [95% CI] | | | | |
| --- | --- | --- | --- | --- | --- |
|  | Stable normal BMI, n (%) 1,802 (48) | Persistent low BMI, n (%) 1,062 (28) | Early overweight resolving, n (%) 303 (8) | Childhood onset obesity, n (%) 164 (5) | Adolescent onset overweight, n (%) 455 (12) |
| **DEMOGRAPHIC AND SOCIOECONOMIC** | | | | | |
| **Sex**, ref: Male |  |  |  |  |  |
| Female | ref | 1.37 [1.16–1.62] | 0.91 [0.72–1.17] | 1.29 [0.96–1.74] | 1.39 [1.15–1.68] |
| **Birth year**, ref: 1993–1997 |  |  |  |  |  |
| 1985–1989 | ref | 0.95 [0.68–1.35] | 0.77 [0.47–1.25] | 0.60 [0.29–1.23] | 0.81 [0.55–1.21] |
| **Ethnicity**, ref: White |  |  |  |  |  |
| South Asian | ref | 1.69 [1.33–2.15] | 0.96 [0.65–1.43] | 1.66 [1.08–2.53] | 1.29 [0.98–1.71] |
| **Parental education^1^**, ref: Compulsory education |  |  |  |  |  |
| Further education | ref | 1.06 [0.87–1.29] | 0.89 [0.67–1.17] | 0.96 [0.69–1.35] | 0.96 [0.77–1.19] |
| Higher education | ref | 1.00 [0.79–1.27] | 0.93 [0.66–1.30] | 0.60 [0.38–0.93] | 1.00 [0.77–1.31] |
| **Degree of urbanisation^2^**, ref: Inner city |  |  |  |  |  |
| Suburbs and rural | ref | 0.83 [0.67–1.01] | 0.90 [0.68–1.19] | 0.84 [0.59–1.20] | 0.87 [0.70–1.10] |
| **Townsend quintile^3^**, ref: 1 (low deprivation) |  |  |  |  |  |
| 2 | ref | 0.93 [0.72–1.20] | 0.97 [0.68–1.38] | 1.50 [0.92–2.43] | 1.01 [0.76–1.35] |
| 3 | ref | 0.95 [0.72–1.24] | 1.26 [0.88–1.81] | 1.42 [0.85–2.38] | 0.98 [0.72–1.34] |
| 4 | ref | 1.06 [0.79–1.41] | 0.90 [0.58–1.38] | 1.66 [0.97–2.84] | 1.05 [0.75–1.46] |
| 5 (high deprivation) | ref | 0.86 [0.64–1.17] | 0.91 [0.58–1.42] | 1.61 [0.92–2.82] | 1.13 [0.80–1.59] |
| **PERINATAL** | | | | | |
| **Birth order**, ref: First child |  |  |  |  |  |
| Second child | ref | 0.92 [0.75–1.11] | 0.81 [0.61–1.08] | 1.07 [0.76–1.51] | 0.88 [0.71–1.10] |
| Third or later child | ref | 0.96 [0.75–1.22] | 0.90 [0.64–1.27] | 0.83 [0.53–1.28] | 0.88 [0.67–1.16] |
| **Age of mother at birth (years)** |  |  |  |  |  |
| Per 1 SD increase: 5.3 years | ref | 0.98 [0.89–1.08] | 0.98 [0.86–1.13] | 1.09 [0.92–1.29] | 0.99 [0.89–1.11] |
| **Gestational age (weeks)** |  |  |  |  |  |
| Per 1 SD increase: 1.9 weeks | ref | 1.24 [1.12–1.37] | 0.93 [0.79–1.09] | 0.89 [0.73–1.07] | 1.10 [0.98–1.24] |
| **Birth weight (grams)** |  |  |  |  |  |
| Per 1 SD increase: 565 grams | ref | 0.61 [0.55–0.68] | 1.46 [1.25–1.70] | 1.22 [1.01–1.48] | 0.78 [0.69–0.88] |

Presented are adjusted odds ratios with 95% confidence intervals from multivariable multinomial logistic regression. We adjusted the effect of each exposure for all other demographic, socioeconomic, and perinatal factors. This model included n = 3,786 participants, because of missing data in parental education, birth order, age of mother at birth, gestational age, and birth weight. We took the uncertainty of class membership into account by weighting each participant’s contribution to the regression model by their posterior probability of membership for each of the five BMI trajectories. We used inverse probability weighting to reduce selection bias arising from differences between participants included in the BMI trajectory analysis (vs excluded), see eSupplement Methods.
^1^Parental education based on the age at completion of full-time education, taking the higher value of either parent. Compulsory education is ≤16 years of age; further education is 17–19 years; higher education is ≥20 years. ^2^Degree of urbanisation: inner city includes postcodes LE1–LE5. ^3^Townsend index of socioeconomic deprivation for the year 2001. See eSupplement-Table 2 for coding of questionnaire variables.
Abbreviations: aOR—adjusted odds ratio; BMI—body mass index; CI—confidence interval; SD—standard deviation.

**eSupplement-Table 10: Identification of risk factors: Adjusted associations of demographic, socioeconomic, perinatal, and lifestyle factors with BMI trajectories in participants of the Leicester Respiratory Cohorts.**

|  | **Multivariable model with outcome BMI trajectory**, aOR [95% CI] | | | | |
| --- | --- | --- | --- | --- | --- |
|  | Stable normal BMI, n (%) 1,203 (46) | Persistent low BMI, n (%) 681 (26) | Early overweight resolving, n (%) 208 (8) | Childhood onset obesity, n (%) 134 (5) | Adolescent onset overweight, n (%) 394 (15) |
| **DEMOGRAPHIC AND SOCIOECONOMIC** | | | | | |
| **Sex**, ref: Male |  |  |  |  |  |
| Female | ref | 1.25 [1.02–1.53] | 1.04 [0.77–1.40] | 1.33 [0.92–1.91] | 1.31 [1.04–1.64] |
| **Birth year**, ref: 1993–1997 |  |  |  |  |  |
| 1985–1989 | ref | 1.27 [0.66–2.46] | 1.28 [0.54–3.02] | 0.75 [0.18–3.05] | 0.70 [0.29–1.68] |
| **Ethnicity**, ref: White |  |  |  |  |  |
| South Asian | ref | 1.51 [1.11–2.05] | 1.02 [0.62–1.71] | 2.15 [1.24–3.72] | 1.29 [0.91–1.84] |
| **Parental education^1^**, ref: Compulsory education |  |  |  |  |  |
| Further education | ref | 1.00 [0.79–1.27] | 0.85 [0.60–1.19] | 1.08 [0.72–1.62] | 0.97 [0.74–1.26] |
| Higher education | ref | 0.98 [0.74–1.30] | 0.90 [0.60–1.35] | 0.59 [0.34–1.03] | 1.11 [0.81–1.52] |
| **Degree of urbanisation^2^**, ref: Inner city |  |  |  |  |  |
| Suburbs and rural | ref | 0.78 [0.61–1.00] | 0.88 [0.63–1.23] | 0.79 [0.51–1.23] | 0.91 [0.69–1.19] |
| **Townsend quintile^3^**, ref: 1 (low deprivation) |  |  |  |  |  |
| 2 | ref | 0.87 [0.65–1.17] | 1.03 [0.67–1.57] | 1.50 [0.85–2.66] | 1.00 [0.72–1.39] |
| 3 | ref | 0.91 [0.66–1.26] | 1.33 [0.86–2.05] | 1.37 [0.74–2.53] | 0.94 [0.65–1.34] |
| 4 | ref | 1.04 [0.73–1.47] | 0.95 [0.56–1.60] | 1.47 [0.77–2.82] | 1.06 [0.71–1.57] |
| 5 (high deprivation) | ref | 0.82 [0.56–1.21] | 1.02 [0.58–1.79] | 1.51 [0.76–3.00] | 1.12 [0.73–1.71] |
| **PERINATAL** | | | | | |
| **Birth order**, ref: First child |  |  |  |  |  |
| Second child | ref | 0.93 [0.74–1.18] | 0.77 [0.55–1.09] | 1.09 [0.72–1.66] | 0.84 [0.65–1.09] |
| Third or later child | ref | 1.03 [0.77–1.37] | 0.77 [0.51–1.16] | 0.70 [0.41–1.20] | 0.77 [0.55–1.06] |
| **Age of mother at birth (years)** |  |  |  |  |  |
| Per 1 SD increase: 5.3 years | ref | 0.96 [0.86–1.09] | 1.03 [0.87–1.22] | 1.20 [0.97–1.47] | 1.00 [0.87–1.14] |
| **Gestational age (weeks)** |  |  |  |  |  |
| Per 1 SD increase: 1.9 weeks | ref | 1.27 [1.12–1.43] | 0.88 [0.73–1.07] | 0.82 [0.66–1.02] | 1.10 [0.96–1.26] |
| **Birth weight (grams)** |  |  |  |  |  |
| Per 1 SD increase: 565 grams | ref | 0.61 [0.53–0.70] | 1.53 [1.27–1.84] | 1.33 [1.06–1.67] | 0.80 [0.68–0.92] |
| **LIFESTYLE** | | | | | |
| **Smoking during pregnancy**, ref: No |  |  |  |  |  |
| Yes | ref | 0.64 [0.45–0.91] | 1.22 [0.79–1.88] | 1.50 [0.88–2.54] | 0.97 [0.68–1.40] |
| **Breastfeeding**, ref: Yes |  |  |  |  |  |
| No | ref | 1.06 [0.85–1.33] | 1.04 [0.75–1.44] | 1.56 [1.07–2.29] | 1.17 [0.91–1.50] |
| **Secondhand tobacco smoke^4^**, ref: No |  |  |  |  |  |
| Yes | ref | 0.94 [0.74–1.18] | 1.37 [0.98–1.91] | 1.22 [0.81–1.85] | 1.15 [0.89–1.49] |
| **Vigorous physical activity^5^**, ref: 0–3 hours per week |  |  |  |  |  |
| ≥4 hours per week | ref | 1.04 [0.85–1.29] | 0.86 [0.64–1.17] | 0.64 [0.44–0.93] | 0.75 [0.59–0.94] |
| **Fresh fruits intake^6^**, ref: Once daily or more |  |  |  |  |  |
| Less than once daily | ref | 1.09 [0.86–1.38] | 1.03 [0.73–1.44] | 0.98 [0.64–1.50] | 1.19 [0.92–1.54] |

Presented are adjusted odds ratios with 95% confidence intervals from multinomial logistic regression. We adjusted the effect of each exposure for all other demographic, socioeconomic, perinatal, and lifestyle factors in this table; we did not include all detailed dietary intake variables because of too much missing data. This model included n = 2,620 participants, because of missing data in exposure variables. We recoded breastfeeding and physical activity to avoid small categories. We did not include other detailed dietary intake because of >50% missing data. We took the uncertainty of class membership into account by weighting each participant’s contribution to the regression model by their posterior probability of membership for each of the five BMI trajectories. We used inverse probability weighting to reduce selection bias arising from differences between participants included in the BMI trajectory analysis (vs excluded), see eSupplement Methods.
^1^Parental education based on the age at completion of full-time education, taking the higher value of either parent. Compulsory education is ≤16 years of age; further education is 17–19 years; higher education is ≥20 years. ^2^Degree of urbanisation: inner city includes postcodes LE1–LE5. ^3^Townsend index of socioeconomic deprivation for the year 2001. ^4^Secondhand tobacco smoke exposure between 0–18 years due to smoking of household member. ^5^Vigorous physical activity: average of a participant’s answers at ages 6–18 years. ^6^Dietary intake: average of a participant’s answers at ages 4–18 years. See eSupplement Materials for coding of questionnaire variables.
Abbreviations: aOR—adjusted odds ratio; BMI—body mass index; CI—confidence interval; SD—standard deviation.

**eSupplement-Table 11: Identification of risk factors: Adjusted associations of demographic, socioeconomic, perinatal, and lifestyle factors with BMI trajectories in South Asian participants of the Leicester Respiratory Cohorts.**

|  | **Multivariable model with outcome BMI trajectory**, aOR [95% CI] | | | | |
| --- | --- | --- | --- | --- | --- |
|  | Stable normal BMI, n (%) 242 (34) | Persistent low BMI, n (%) 265 (37) | Early overweight resolving, n (%) 31 (4) | Childhood onset obesity, n (%) 46 (6) | Adolescent onset overweight, n (%) 124 (18) |
| **DEMOGRAPHIC AND SOCIOECONOMIC** | | | | | |
| **Sex**, ref: Male |  |  |  |  |  |
| Female | ref | 1.45 [0.94–2.23] | 1.33 [0.58–3.05] | 0.85 [0.41–1.79] | 1.00 [0.61–1.66] |
| **Parental education^1^**, ref: Higher education |  |  |  |  |  |
| Further education | ref | 0.82 [0.51–1.33] | 1.37 [0.54–3.47] | 1.20 [0.53–2.73] | 0.68 [0.39–1.19] |
| Compulsory education | ref | 0.69 [0.39–1.23] | 1.04 [0.33–3.24] | 0.82 [0.30–2.23] | 0.53 [0.27–1.04] |
| **Townsend quintile^2^**, ref: 5 (high deprivation) |  |  |  |  |  |
| 4 | ref | 0.87 [0.51–1.47] | 0.74 [0.26–2.06] | 0.84 [0.34–2.03] | 0.67 [0.36–1.24] |
| 3 | ref | 0.67 [0.36–1.27] | 1.12 [0.37–3.39] | 0.81 [0.28–2.34] | 0.69 [0.34–1.41] |
| 2 | ref | 0.74 [0.34–1.59] | 0.77 [0.16–3.79] | 0.70 [0.17–2.90] | 0.62 [0.25–1.51] |
| 1 (low deprivation) | ref | 1.43 [0.57–3.63] | 0.56 [0.07–4.39] | 0.94 [0.18–4.83] | 0.80 [0.26–2.46] |
| **PERINATAL** | | | | | |
| **Birth order**, ref: First child |  |  |  |  |  |
| Second child | ref | 0.64 [0.38–1.08] | 0.26 [0.10–0.73] | 0.66 [0.26–1.66] | 0.69 [0.38–1.24] |
| Third or later child | ref | 0.67 [0.36–1.25] | 0.35 [0.12–1.04] | 0.79 [0.28–2.26] | 0.55 [0.26–1.14] |
| **Age of mother at birth (years)** |  |  |  |  |  |
| Per 1 SD increase: 5.3 years | ref | 1.16 [0.89–1.50] | 1.30 [0.83–2.02] | 1.31 [0.86–2.01] | 1.10 [0.82–1.48] |
| **Gestational age (weeks)** |  |  |  |  |  |
| Per 1 SD increase: 1.9 weeks | ref | 1.34 [1.03–1.75] | 0.80 [0.46–1.39] | 1.16 [0.70–1.91] | 1.27 [0.93–1.74] |
| **Birth weight (grams)** |  |  |  |  |  |
| Per 1 SD increase: 565 grams | ref | 0.56 [0.42–0.74] | 1.95 [1.18–3.24] | 1.40 [0.88–2.21] | 0.71 [0.52–0.99] |
| **LIFESTYLE** | | | | | |
| **Breastfeeding**, ref: Yes |  |  |  |  |  |
| No | ref | 1.18 [0.72–1.95] | 0.81 [0.29–2.30] | 1.31 [0.57–2.98] | 0.87 [0.48–1.60] |
| **Secondhand tobacco smoke^3^**, ref: No |  |  |  |  |  |
| Yes | ref | 0.59 [0.37–0.94] | 1.20 [0.51–2.81] | 0.84 [0.38–1.85] | 0.89 [0.52–1.50] |
| **Vigorous physical activity^4^**, ref: 0–3 hours per week |  |  |  |  |  |
| ≥4 hours per week | ref | 0.98 [0.63–1.53] | 1.10 [0.47–2.57] | 0.52 [0.23–1.15] | 0.97 [0.58–1.61] |
| **Fresh fruits intake^5^**, ref: Once daily or more |  |  |  |  |  |
| Less than once daily | ref | 1.05 [0.65–1.69] | 0.75 [0.28–1.99] | 0.91 [0.39–2.09] | 0.94 [0.53–1.65] |

Presented are adjusted odds ratios with 95% confidence intervals from multinomial logistic regression. We adjusted the effect of each exposure for all other demographic, socioeconomic, perinatal, and lifestyle factors in this table; we did not include all detailed dietary intake variables because of too much missing data. This model included n = 708 participants, because of missing data in exposures. We recoded breastfeeding and physical activity to avoid small categories. We did not include birth year because all South Asian participants were from the newer birth years (1993–1997). We did not include degree of urbanisation because >90% lived in the inner city. We did not include smoking during pregnancy because <1% was exposed. We did not include other detailed dietary intake because of >50% missing data. We took the uncertainty of class membership into account by weighting each participant’s contribution to the regression model by their posterior probability of membership for each of the five BMI trajectories. We used inverse probability weighting to reduce selection bias arising from differences between participants included in the BMI trajectory analysis (vs excluded), see eSupplement Methods.
^1^Parental education based on the age at completion of full-time education, taking the higher value of either parent. Compulsory education is ≤16 years of age; further education is 17–19 years; higher education is ≥20 years. ^2^Townsend index of socioeconomic deprivation for the year 2001. ^3^Secondhand tobacco smoke exposure between 0–18 years due to smoking of household member. ^4^Vigorous physical activity: average of a participant’s answers at ages 6–18 years. ^5^Dietary intake: average of a participant’s answers at ages 4–18 years. See eSupplement-Table 2 for coding of questionnaire variables.
Abbreviations: aOR—adjusted odds ratio; BMI—Body mass index; CI—Confidence interval; SD—standard deviation.

**eSupplement-Table 12: Identification of risk factors: Adjusted associations of demographic, socioeconomic, perinatal, and lifestyle factors with BMI trajectories in White participants of the Leicester Respiratory Cohorts.**

|  | **Multivariable model with outcome BMI trajectory**, aOR [95% CI] | | | | |
| --- | --- | --- | --- | --- | --- |
|  | Stable normal BMI, n (%) 969 (50) | Persistent low BMI, n (%) 430 (22) | Early overweight resolving, n (%) 177 (9) | Childhood onset obesity, n (%) 91 (5) | Adolescent onset overweight, n (%) 277 (14) |
| **DEMOGRAPHIC AND SOCIOECONOMIC** | | | | | |
| **Sex**, ref: Male |  |  |  |  |  |
| Female | ref | 1.17 [0.93–1.48] | 0.99 [0.72–1.36] | 1.51 [0.99–2.29] | 1.41 [1.10–1.83] |
| **Birth year**, ref: 1993–1997 |  |  |  |  |  |
| 1985–1989 | ref | 1.26 [0.65–2.43] | 1.21 [0.52–2.86] | 0.74 [0.18–3.01] | 0.69 [0.29–1.65] |
| **Parental education^1^**, ref: Compulsory education |  |  |  |  |  |
| Further education | ref | 0.99 [0.76–1.29] | 0.8 [0.55–1.15] | 1.07 [0.68–1.68] | 0.92 [0.69–1.23] |
| Higher education | ref | 0.86 [0.61–1.20] | 0.92 [0.59–1.44] | 0.45 [0.21–0.95] | 0.96 [0.67–1.39] |
| **Degree of urbanisation^2^**, ref: Inner city |  |  |  |  |  |
| Suburbs and rural | ref | 0.74 [0.57–0.96] | 0.86 [0.61–1.22] | 0.75 [0.47–1.18] | 0.89 [0.67–1.18] |
| **Townsend quintile^3^**, ref: 1 (low deprivation) |  |  |  |  |  |
| 2 | ref | 0.89 [0.66–1.22] | 1.02 [0.67–1.57] | 1.59 [0.87–2.89] | 0.99 [0.70–1.39] |
| 3 | ref | 1.00 [0.71–1.40] | 1.28 [0.82–2.02] | 1.39 [0.72–2.72] | 0.90 [0.61–1.32] |
| 4 | ref | 1.12 [0.75–1.65] | 0.92 [0.52–1.63] | 1.58 [0.76–3.27] | 1.10 [0.71–1.70] |
| 5 (high deprivation) | ref | 0.68 [0.42–1.10] | 0.89 [0.47–1.67] | 1.37 [0.61–3.05] | 0.95 [0.58–1.58] |
| **PERINATAL** | | | | | |
| **Birth order**, ref: First child |  |  |  |  |  |
| Second child | ref | 1.02 [0.78–1.33] | 0.90 [0.63–1.30] | 1.20 [0.75–1.91] | 0.84 [0.63–1.13] |
| Third or later child | ref | 1.17 [0.84–1.62] | 0.87 [0.55–1.36] | 0.60 [0.31–1.14] | 0.83 [0.57–1.20] |
| **Age of mother at birth (years)** |  |  |  |  |  |
| Per 1 SD increase: 5.3 years | ref | 0.94 [0.81–1.08] | 0.99 [0.82–1.20] | 1.20 [0.94–1.54] | 1.01 [0.86–1.17] |
| **Gestational age (weeks)** |  |  |  |  |  |
| Per 1 SD increase: 1.9 weeks | ref | 1.27 [1.10–1.45] | 0.91 [0.74–1.11] | 0.78 [0.62–0.99] | 1.09 [0.94–1.27] |
| **Birth weight (grams)** |  |  |  |  |  |
| Per 1 SD increase: 565 grams | ref | 0.63 [0.54–0.73] | 1.45 [1.19–1.76] | 1.29 [1.00–1.67] | 0.80 [0.68–0.95] |
| **LIFESTYLE** | | | | | |
| **Smoking during pregnancy**, ref: No |  |  |  |  |  |
| Yes | ref | 0.60 [0.42–0.88] | 1.16 [0.74–1.82] | 1.32 [0.76–2.32] | 0.91 [0.62–1.32] |
| **Breastfeeding**, ref: Yes |  |  |  |  |  |
| No | ref | 0.98 [0.76–1.27] | 1.05 [0.74–1.47] | 1.56 [1.01–2.41] | 1.18 [0.89–1.55] |
| **Secondhand tobacco smoke^4^**, ref: No |  |  |  |  |  |
| Yes | ref | 1.05 [0.81–1.38] | 1.41 [0.98–2.02] | 1.30 [0.80–2.09] | 1.21 [0.90–1.63] |
| **Vigorous physical activity^5^**, ref: 0–3 hours per week |  |  |  |  |  |
| ≥4 hours per week | ref | 1.09 [0.85–1.39] | 0.82 [0.60–1.13] | 0.67 [0.44–1.01] | 0.70 [0.54–0.90] |
| **Fresh fruits intake^6^**, ref: Once daily or more |  |  |  |  |  |
| Less than once daily | ref | 1.08 [0.82–1.42] | 1.06 [0.74–1.53] | 0.95 [0.58–1.55] | 1.27 [0.95–1.70] |

Presented are adjusted odds ratios with 95% confidence intervals from multinomial logistic regression. We adjusted the effect of each exposure for all other demographic, socioeconomic, perinatal, and lifestyle factors in this table; we did not include all detailed dietary intake variables because of too much missing data. This model included n = 1,944 participants, because of missing data in exposures. We recoded breastfeeding and physical activity to avoid small categories. We did not include other detailed dietary intake because of >50% missing data. We took the uncertainty of class membership into account by weighting each participant’s contribution to the regression model by their posterior probability of membership for each of the five BMI trajectories. We used inverse probability weighting to reduce selection bias arising from differences between participants included in the BMI trajectory analysis (vs excluded), see eSupplement Methods.
^1^Parental education based on the age at completion of full-time education, taking the higher value of either parent. Compulsory education is ≤16 years of age; further education is 17–19 years; higher education is ≥20 years. ^2^Degree of urbanisation: inner city includes postcodes LE1–LE5. ^3^Townsend index of socioeconomic deprivation for the year 2001. ^4^Secondhand tobacco smoke exposure between 0–18 years due to smoking of household member. ^5^Vigorous physical activity: average of a participant’s answers at ages 6–18 years. ^6^Dietary intake: average of a participant’s answers at ages 4–18 years. See eSupplement-Table 2 for coding of questionnaire variables.
Abbreviations: aOR—adjusted odds ratio; BMI—body mass index; CI—confidence interval; SD—standard deviation.
