## Supplementary material for "Ethnic and Social Health Inequalities in Body Mass Index Trajectories through Childhood and Adolescence: A Longitudinal Population-Based Study in Leicestershire UK": GRoLTS checklist

| **Checklist Item** | **Reported?** |  |
| --- | --- | --- |
| 1. | Is the metric of time used in the statistical model reported? | Yes  page 6  Figure 2 |
| 2. | Is information presented about the mean and variance of time within a wave? | Partly: No use of discrete waves  eTables 5&6 |
| 3a. | Is the missing data mechanism reported? | Yes pages 6&7  eFigure 1  eMethods |
| 3b. | Is a description provided of what variables are related to attrition/missing data? | Yes  Tables 1&2  eFigures 1&2  eTable 3 |
| 3c. | Is a description provided of how missing data in the analyses were dealt with? | Yes  Page 7  eMethods  eFigure 1 |
| 4. | Is information about the distribution of the observed variables included? | Yes  Figure 2 |
| 5. | Is the software mentioned? | Yes  Page 7,  eMethods |
| 6a. | Are alternative specifications of within-class heterogeneity considered (e.g., LGCA vs. LGMM) and clearly documented? If not, was sufficient justification provided as to eliminate certain specifications from consideration? | Yes: considered and justified to eliminate GMM  Page 6,  eMethods |
| 6b. | Are alternative specifications of the between-class differences in variance–covariance matrix structure considered and clearly documented? If not, was sufficient justification provided as to eliminate certain specifications from consideration? | Yes: considered and justified to set equal matrix eMethods |
| 7. | Are alternative shape/functional forms of the trajectories described? | Yes: considered polynomials and splines  Page 6  eMethods |
| 8. | If covariates have been used, can analyses still be replicated? | No covariates used in GBTM |
| 9. | Is information reported about the number of random start values and final iterations included? | Yes  Page 7  eMethods |
| 10. | Are the model comparison (and selection) tools described from a statistical perspective? | Yes Page 7  eMethods |
| 11. | Are the total number of fitted models reported, including a one-class solution? | Yes Page 7  eMethods |
| 12. | Are the number of cases per class reported for each model (absolute sample size, or proportion)? | Yes  eTable 7 |
| 13. | If classification of cases in a trajectory is the goal, is entropy reported? | Yes  eTable 7 |
| 14a. | Is a plot included with the estimated mean trajectories of the final solution? | Yes  Figure 2 |
| 14b. | Are plots included with the estimated mean trajectories for each model? | Yes  eFigures 3&4 |
| 14c. | Is a plot included of the combination of estimated means of the final model and the observed individual trajectories split out for each latent class? | Yes  Figure 2 |
| 15. | Are characteristics of the final class solution numerically described (i.e., means, *SD/SE, n*, CI, etc.)? | Yes  Figure 2 with mean and CIs  No Table |
| 16. | Are the syntax files available (either in the appendix, supplementary materials, or from the authors)? | Yes on GitHub |
